# Epigenetic factors in the 22q11.2 deletion syndrome in relation to stress and schizophrenia

**DOI:** 10.1101/2024.06.23.24309352

**Authors:** Chuan Jiao, Fanny Demars, Anton Iftimovici, Qin He, Oussama Kebir, Anushree Tripathi, Hugo Turbé, Caroline Demily, Marie-Odile Krebs, Thérèse M Jay, Boris Chaumette

## Abstract

22q11.2 deletion syndrome (22q11.2DS) stands out as one of the most significant risk factors for schizophrenia (SCZ), with approximately 40% of individuals with 22q11.2DS experiencing psychosis. The presence of discordant phenotypes among monozygotic twins, along with the involvement of environmental factors in the multiple-hit model hypothesis for psychosis onset, underscores the potential role of epigenetic modifications in the development of neuropsychiatric disorders among individuals with 22q11.2DS. To gain a deeper understanding of the underlying biological mechanisms, we conducted a translational study using three datasets: a genome-wide methylation dataset from peripheral blood of individuals with 22q11.2DS with or without SCZ, a microRNA expression dataset from the same cohort, and a second genome-wide methylation dataset obtained from a mouse model exploring gene-environment interactions. Human recruitment was carried out at a specialized center focusing on rare psychiatric disorders and included one pair of monozygotic twins discordant for SCZ. In the animal model, DNA extraction was performed from the prefrontal cortex among four groups : wild-type and Df(h22q11)/+ mice, with or without exposure to acute stress. This study identified alterations in DNA methylation and microRNA expression linked to the 22q11.2 deletion as well as SCZ within the context of the deletion in humans. The results were then compared to the effects of the corresponding deletion and stress in the mouse model. Notably, four genes (*ZBTB20, SHANK3, GRAMD1B, XKR4*) overlapped across all comparisons. Pathway analysis evealed epigenetic differences in the Wnt pathway associated with stress and SCZ within the context of the deletion. These findings support the hypothesis that the onset of SCZ in individuals with 22q11.2DS may be influenced by epigenetic mechanisms, both within and outside the implicated region, under the influence of environmental stressors. If replicated, these findings could be used to develop biomarkers for early diagnosis in del22q11 carriers and to explore new targeted therapeutic strategies.

## Introduction

Schizophrenia (SCZ) is a frequent severe psychotic disorder resulting from complex interactions between genetic factors and pre/postnatal exposure to environmental risk factors. One of the highest genetic risk factors for SCZ is the deletion of the 22q11.2 chromosomal region (odds ratio=67.7) [1], affecting 1 children over 2000-4000 live births worldwide [2, 3]. About 40% of individuals with the 22q11.2 deletion syndrome (22q11.2DS, also called DiGeorge Syndrome) experience psychosis in their adulthood [4] and the deletion could account for up to 0.5-1% of all SCZ cases [5] The typical deletion is 3 Mb long, producing a haploinsufficiency of ∼90 genes [6]. The genetic-phenotype correlation remained challenging, patients with identical deletions exhibiting diverse clinical phenotypes. Thus, haploinsufficiency does not fully explain the varying penetrance and severity of clinical phenotypes in different patients [7, 8]. Identifying other pathophysiological factors that could modify the development of psychosis in 22q11.2DS behind the deletion itself is therefore crucial. Possible additional genetic hits, for instance other Copy Number Variants, could mediate this variable penetrance [9]. But, no common or rare variants within the 22q11.2 region were significantly associated with SCZ [10]. Finally, polygenic risk scores for schizophrenia have been reported to be higher in 22q11.2DS individuals with schizophrenia than in 22q11.2DS without SCZ [10, 11].

However, the existence of discordant phenotypes in monozygotic twins supports the involvement of epigenetic modifications in the onset of neuropsychiatric disorders in 22q11.2DS [7, 12]. DNA methylation differences in blood samples of 22q11.2DS individuals with/without SCZ have been found in imprinted genes and MHC locus [13]. An episignature of the 22q11.2DS has been reported: this classifier, built on 160 CpGs could discriminate 22q11.2DS and healthy controls, regardless their psychiatric status [14]. DNA methylation analyses have been conducted in 22q11.2DS at birth to test if their methylation pattern is associated with mental disorder later in life [15]. However, since DNA methylation undergoes dynamic changes during life span, it is essential to identify the DNA methylation differences in 22q11.2DS with or without psychiatric diagnosis in adulthood. Indeed, our group has demonstrated that DNA methylation changes could occur during the onset of psychosis [16]. Recently, we have also reported microRNA (miRNA) expression changes accompanying the emergence of psychosis in adolescence [17]. Interestingly, miRNA maturation requires the expression of DGCR8, a protein encoded by the gene *DGCR8* (DiGeorge syndrome Critical Region 8) located in the 22q11.2 region. It binds to Drosha allowing for cleaving the primary miRNAs into precursor miRNAs [18]. Due to the haploinsufficiency of the DGCR8 gene in patients with 22q11.2DS, imbalanced DGCR8-Drosha protein stoichiometry reduces the biogenesis of hundreds of precursor miRNA. miRNAs are essential post-transcriptional regulators of gene expression and regulates many physiological mechanisms including brain development and functions [18, 19]. Their dysregulation induces behavioral and neuronal deficits [20–22]. miRNA dysregulation has been reported in patients with 22q11.2DS [23–25] and SCZ [26, 27], but it is unclear how miRNA dysregulation in patients with 22q11.2DS may increase the risk of SCZ [28]. Moreover, miR-185, a miRNA known to be associated with SCZ, is located in the 22q11.2 region and its deletion could dysregulate gene expression and ultimately leading to SCZ [29].

In this study, we hypothesize that both DNA methylation and miRNA expression are altered in individuals with 22q11.2DS and differed between those with/without SCZ. We took advantage of a cohort recruited in specialized French clinical settings (Centers for Rare Psychiatric Disorder), including two monozygotic twins, discordant for SCZ.

In line with the multiple-hit model hypothesis of SCZ, we believe that epigenetic changes could be related to environmental factors. The predominant environmental risk factor for 22q11.2DS patients who develop SCZ could be psychosocial stress [30]. Psychosocial stress can involve epigenetic modifications [31] and has been shown to be associated with SCZ onset [32, 33]. Therefore, we aimed to explore the epigenome in the context of stress to understand the biological mechanisms underlying the incomplete penetrance of SCZ in 22q11.2DS. We designed an animal study that facilitates our access to brain tissue, and allows for controlling the genetic background and the exposition to stressful environmental factors [34]. Here, we used a mouse model Df(h22q11)/+ characterized by a hemizygous deletion of the mouse chromosome 16 which has a high degree of conservation with the region of human 22q11.2 microdeletion [35]. The phenotypic features in the mouse model are comparable to the psychiatric and non-psychiatric features of humans with 22q11DS [36]. In particular, we have previously shown that the Df(h22q11)/+ mice exhibit social and sensorimotor processing deficits, as well as impaired cognitive capabilities including working memory, attentional set-shifting, behavioral flexibility, and decision-making [37]. Interestingly, as in humans, evidence supports that the deletion confers vulnerability revealed by additional environmental hits such as environmental stress. We completed the human study, by a genome-wide exploration of DNA methylation in the prefrontal cortex of the Df(h22q11)/+ mice.

The **Figure 1** summarizes the design of this translational study. First, we explored the genome-wide DNA methylation and miRNA expression differences between 22q11.2DS patients and controls, and then we compared among 22q11.2DS patients, those with/without SCZ. Notably, a pair of discordant monozygotic 22q11.2DS twins was included. Second, we compare DNA methylation between four groups of mice following a Gene x Environment interaction design: a group of wild-type mice and a group of Df(h22q11)/+ mice, each one being exposed or not to an environmental acute stress.

**Figure 1.**
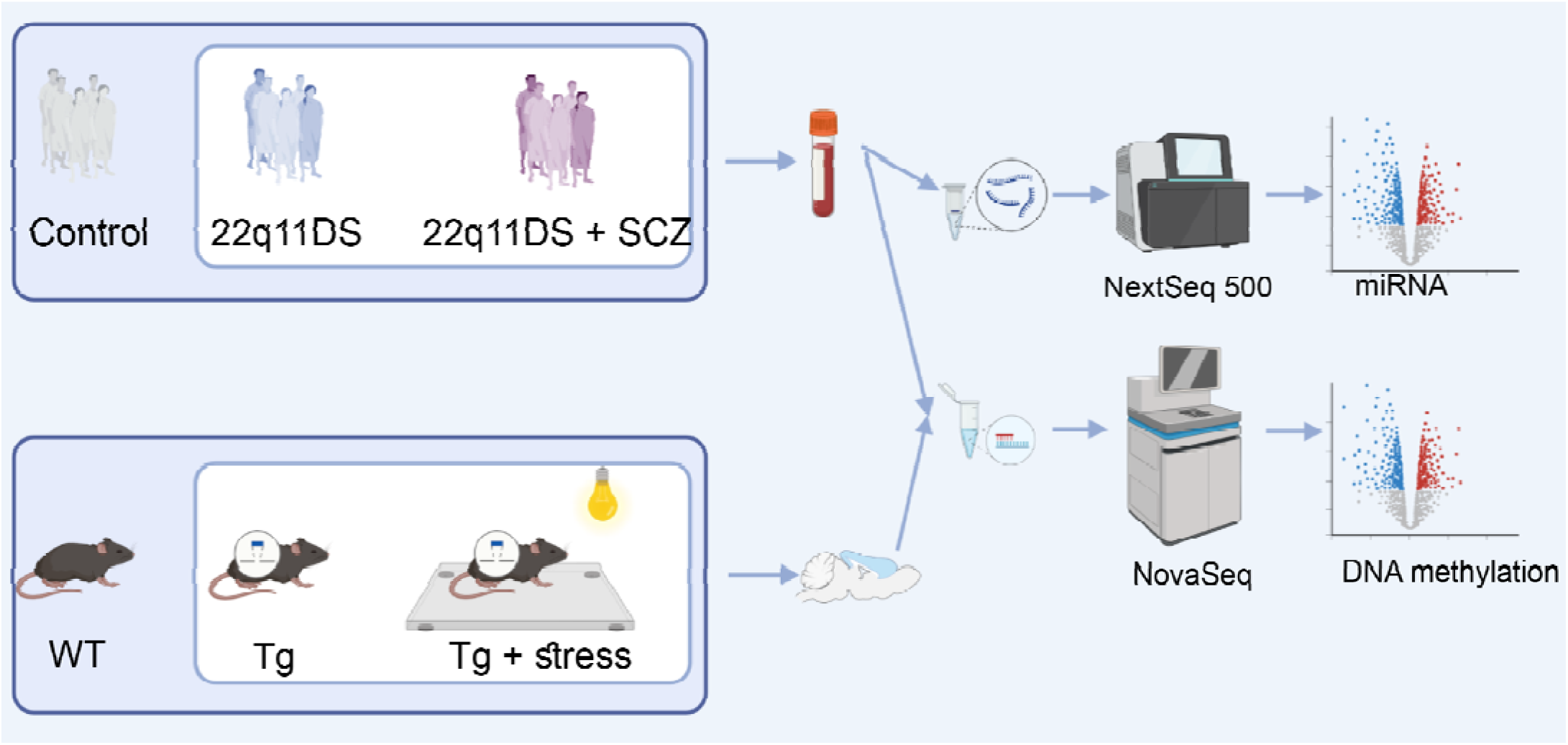
Study design. A methylomic-wide association study (MWAS) was conducted in blood to compare 22q11.2 deletion carriers and non-carriers. Then, among 22q11.2 deletion carriers, we compared those with SCZ to those without SCZ. We obtained genome-wide methylation data from the prefrontal cortex of transgenic (Tg) mice and wild-type (WT) mice. We conducted a MWAS between Tg and WT and, among the Tg, between stressed and non-stressed mice. In addition, we compared the miRNA expression between 22q11.2 carriers and non-carriers as well as, among the carriers, between individuals with and without SCZ. Figure created with BioRender.com.

## Materials and Methods

### 1. Human cohort

The individuals with 22q11DS and non-carriers individuals were recruited at Sainte-Anne Hospital (Paris, France) as part of the PSYDEV collection (“Etude familiale et génétique des aspects développementaux des maladies psychiatriques”) and the ICAAR cohort (“Influence du Cannabis sur l’émergence de symptômes psychopathologiques des Adolescents et jeunes Adultes présentant un état mental à Risque”, 2009–2014). Two monozygotic twins with a discordant diagnosis for SCZ were recruited in another center for Rare Psychiatric Disorders in Lyon (Genopsy).The recruitments were approved by the institutional ethics committee “Comité de protection des personnes, Ile-de-France IV, Paris, France,” and written informed consents were obtained from all participants or their legal representatives when applicable, following the Declaration of Helsinki. A total of 28 individuals were included for the DNA methylation study, and 32 for the miRNA study. All individuals were Caucasian as confirmed by their genotyping data (Infinium PsychArray data analyzed with the software Peddy).

### 2. Animals preparation

All experimental procedures were carried out on 10 adult male transgenic Df(h22q11)/+ mice (Tg) and 10 wild-type littermates (WT) (10–13 weeks of age) obtained from Taconics Biosciences thanks to the IMI NEWMEDS collaboration, as reported in [37]. Half of the animals of each group (5 Tg and 5 WT) were subjected to acute stress, as previously described [38]. Experimental protocols were in accordance with National (JO 887–848) and European (86/609/EEC) legislation regarding animal experimentation. The acute stress procedure was performed by an experimenter blind to the mice’s genotype. Briefly, mice were placed on an unsteady platform (20□cm□×□20□cm) at a height of 1m above the ground in front of a bright light (1500lux) for 30 minutes. Under these conditions, mice show freezing behavior. Four groups of 5 animals were defined: transgenic mice and wild-type mice exposed to stress or not.

### 3. Sample extraction

#### 3.1 DNA and RNA preparation in human

Biological samples were obtained from the Biological Resource Center NSPN of the GHU Paris Psychiatrie & Neurosciences, which is responsible for the centralized management of biological data collection. DNA was extracted from whole blood (EDTA tubes) using a salting-out protocol and then purified with the PureGen Qiagen kit. miRNAs were extracted from PAXgene with miRNeasy Advanced Kit (QIAGEN).

#### 3.2 DNA preparation in mice

Within 30 min of the end of the stress procedure, mice were anesthetized with isoflurane to withdraw blood from the orbital sinus and then decerebrated. The brains were immediately removed, frozen in isopentane for 1-2 min at -50°C, stored at -80°C until sectioning, and then cryo-sectioned (100 μm). The medial prefrontal cortex (mPFC) was micro-punched (anterior-posterior: 1.75-2.1 mm; mediolateral: 0.3 mm, dorsoventral: 1.9-2.1 mm from the bregma, referring to Paxinos and Watson Atlas). DNA was extracted using the AllPrep mini kit (QIAGEN).

### 4. Epigenome-wide analysis

#### 4.1 Bisulfite sequencing

Methylation data were generated by reduction-presentation bisulfite sequencing (RRBS) [39]. Fragments were selected by gel migration after MSP1 digestion of 500 ng of DNA from each sample. Libraries were built using the Premium Reduced Representation Bisulfite Sequencing kit from Diagenode. In humans, 2 × 100bp paired-end reads were sequenced with NovaSeq with bisulfite conversion efficiencies > 98% and at least 70 million reads per sample. In mice, 2 × 75bp paired-end reads were sequenced with NovaSeq, with bisulfite conversion efficiencies > 99.5% and a minimum of 40 million reads per sample.

#### 4.2 Preprocessing

We used Trim Galore (version v0.6.7) to perform quality checks and adapter trimming on raw reads in fastq format. In addition to the 13 bp Illumina standard adapters, we removed 5 bp adapters from their 5’ and 3’ ends to avoid poor quality or bias based on our FastQC report. After trimming, we also removed too short sequences (less than 20bp). Since our RRBS were generated on the NovaSeq platform, we also replaced the default parameter from -- quality to --2colors.

We used Bismark [40](version: v0.23.1dev) to prepare reference genomes, GRCh38 (hg38) for human and GRCm38 (mm10) for mouse. The trimmed fastq files were imported into Bismark for alignment. To achieve better alignment efficiency, we applied local alignment mode with a function of --score-min based on the length of trimmed reads (i.e. -G,90,0 for the human RRBS data, where G indicates the function type as natural log, 90 is the read length, and 0 is the constant term of the function). The parameter coverage2cytosine was used to merge CpGs from both strands and detect the cytosine methylation. In the analysis, we filtered out CpG sites from X and Y chromosomes and CpG sites from mitochondrial DNA. Cell type deconvolution was conducted with methylCC [41] (v1.16.0) (**Supplementary Table S23**).

#### 4.3 Differential analysis

We used DSS [42] and limma[43] to detect significant differentially methylated CpG (DMPs) and then mapped the significant DMPs into differentially methylated regions (DMRs) using DMRcate [44] (v2.16.0). CpGs without coverage in all samples and a coverage less than 10 in 50% samples were excluded. Surrogate variable analyses (sva package v3.40.0) [45] and principal components analysis (PCA) were respectively used to estimate surrogate variables and PCs for unknown covariates. Principal variance component analysis (PVCA) was used to see the contributions of the covariates on the methylation levels, which were then selectively added into the design model. QQ plot and lambda values were checked to identify the fitness of models. FDR correction was used to adjust the p-values for multiple tests. CpGs with FDR-corrected p-value less than 0.05 were set as significant. CpGs were annotated to genes by their nearest transcription site start (edgeR v3.42.4 [46]). DMRs were detected by DMRcate with the default parameters. The significance of DMRs was evaluated by Fisher combined probability based on CpG FDR-corrected p-values.

### 5. RNA analysis

For the twins with 22q11DS, we conducted a RNAseq analysis. The RNAs were extracted from PAXgenes and the samples were depleted for ribosomal RNA. The libraries were carried out with the NEBNext Ultra II Total-RNA-Seq and then sequenced on the Illumina NovaSeq6000 with the following parameters: 100bp of length, paired-end, 160 millions of reads. We used Trimmomatic (v0.39) [47] to remove the adapter and quality trimming, STAR (v2.7.9a) [48] for alignment, and RSEM (v1.3.3) [49] for quantification. We filtered the genes that were not expressed in both of the twins’ samples.

### 6. miRNA analysis

We investigated miRNA expression differences between 11 individuals with 22q11.2DS patients and 20 non-carriers as well as differences between individuals with 22q11.2DS with or without SCZ. QIAseq® miRNA Library Kit (QIAGEN) was used for library preparation, using unique molecular index assignment. Sequencing was performed on the Illumina NextSeq 500 platform at ICM (Institut du cerveau et de la moelle épinière, Hôpital de la Salpêtrière, Paris, France). The results showed an average total read count of 18 million, with 14% of the total reads being UMI-assigned, of which 30% were aligned to 2,479 miRNAs. miRNAs were filtered with a minimum of 5 read counts in at least 25% of the samples. Differentially expressed miRNAs were detected by DESeq2 [50] v1.32.0. We performed Principal variance component analysis (PVCA) to see the contributions of each covariate. Sex, and age were listed in the model design to correct for confounders. We predicted the target genes using miRDB [51]. We only included targets with a prediction score of more than 60 and restricted the analysis to miRNAs included in the FuncMir Collection, and not targeting more than 2000 predicted genes.

### 7. Enrichment analysis

Gene annotation and pathway enrichment analysis on the identified genes were performed using clusterProfiler [53] v4.6.2. P-values were calculated based on the accumulative hypergeometric distribution, and q-values were calculated using the FDR procedure to account for multiple tests. Pathway databases considered for enrichment analysis included Gene Ontology (GO), KEGG Pathway, and WikiPathways. We used Multi-marker Analysis of GenoMic Annotation (MAGMA) [54] as well as FUMA GWAS (https://fuma.ctglab.nl/) GENE2FUNC [55] to test the gene-set enrichment with the genome-wide association studies (GWAS) of SCZ. Tissue type enrichment analysis was performed by TissueEnrich (v1.18.0) [56] with Human Protein Atlas (HPA) as the gene expression reference database.

## Results

### Differences in DNA methylation associated with the 22q11.2 deletion

To test the effect of the 22q11 deletion on DNA methylation, we compared 14 22q11.2DS patients (age: 26.4 ± 9.3, F/M: 6/8) to 14 individuals without the deletion and matched for age, sex and ancestry (**Table S1**). We detected 1,422 significant DMPs located on 1,165 genes (72.02% of protein-coding genes, **Figure 2** & **Table S2**), and 11 significant DMRs located on 6 genes (**Figure 3**, **Figure S1, Table S4**).

**Figure 2.**
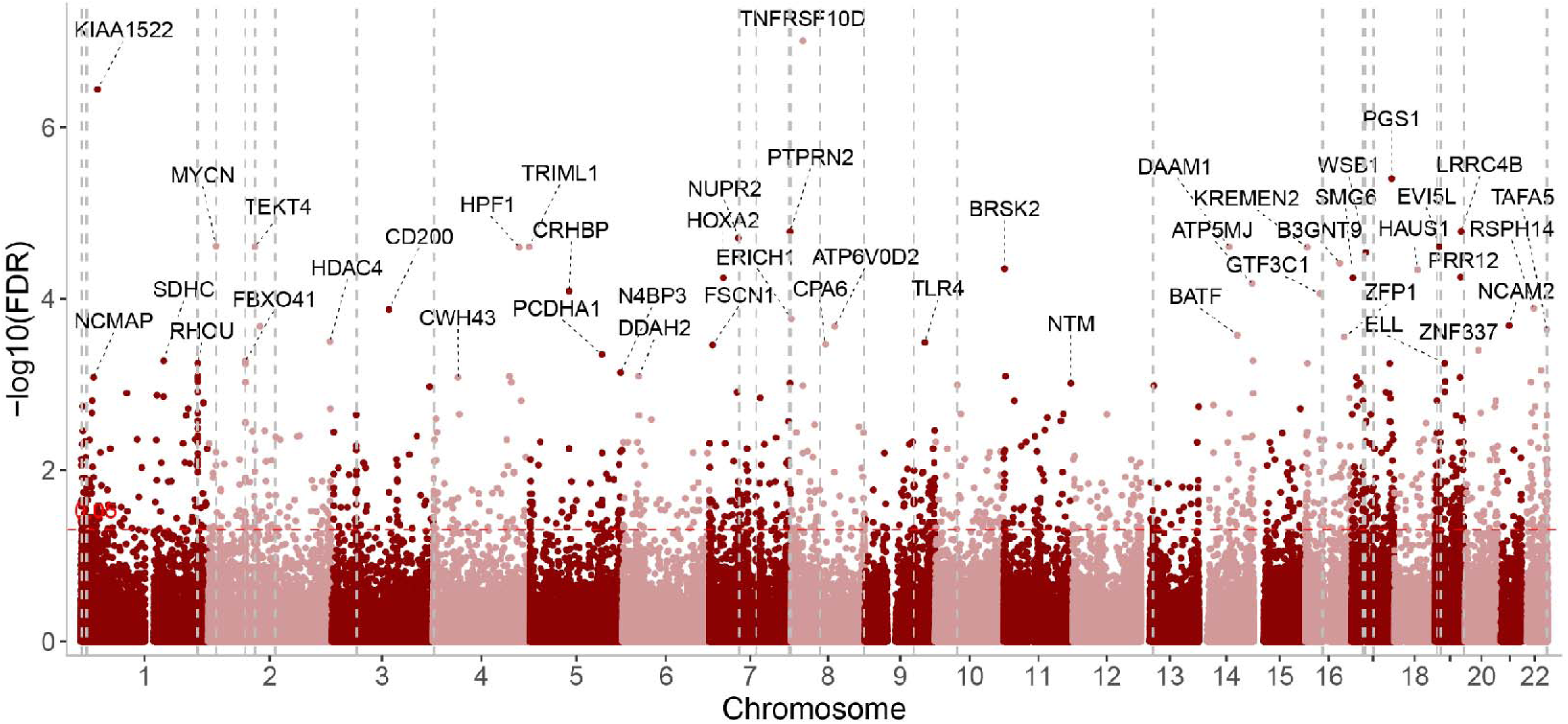
22q11.2 deletion effects on DNA methylation. Manhattan plot of 22q11.2 deletion effects on DNA methylation in humans obtained by comparing 22q11.2 deletion carriers and non-carriers. Gene names were marked when FDR<0.001. The vertical dashed lines show their localization on the genome.. The horizontal red line shows the nominal significance (uncorrected p-value <0.05).

**Figure 3.**
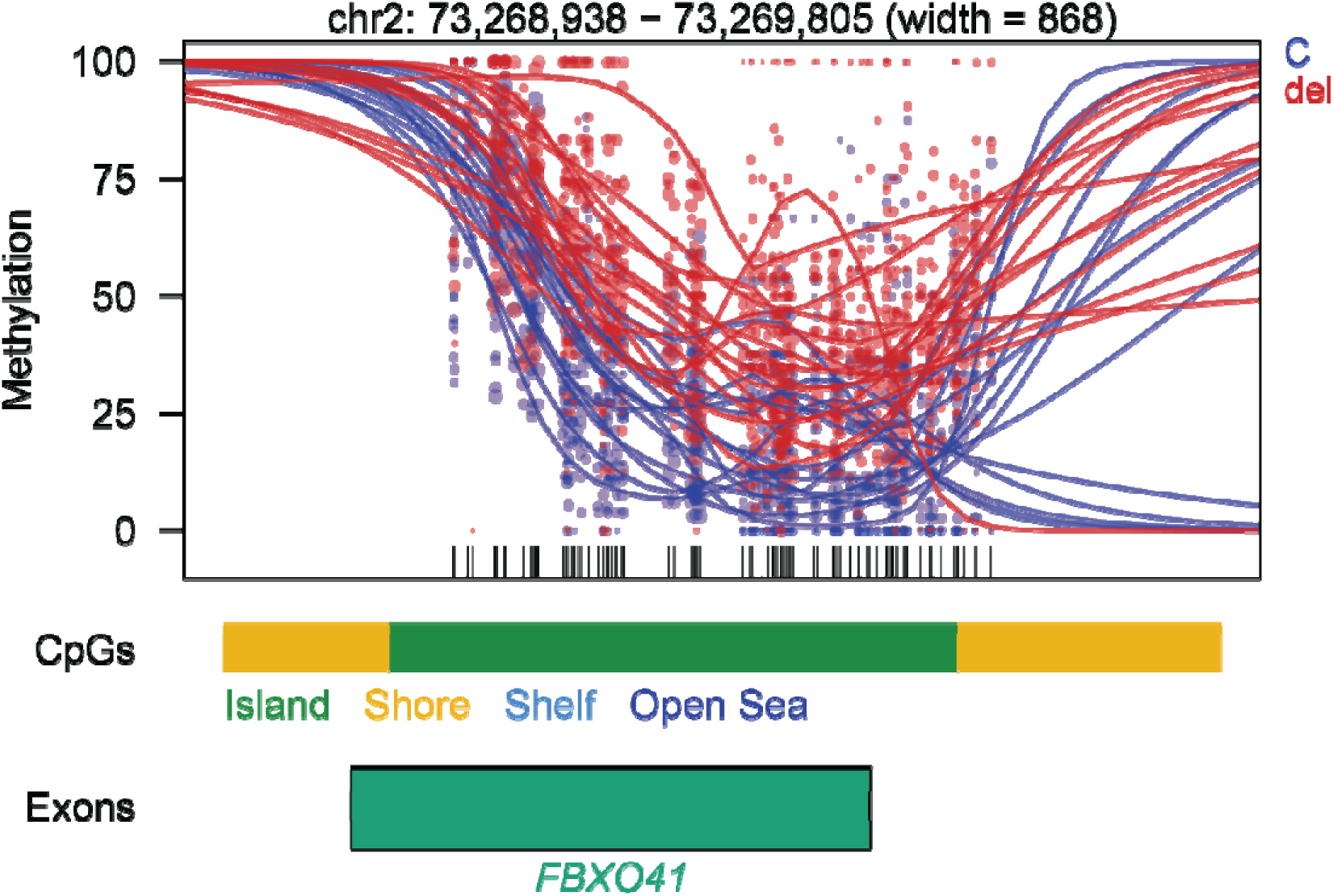
Example of one DMR located in the gene FBXO41. Each dot represents the methylation level of an individual CpG in a single sample, where the size of the dot represents coverage, and the color of the dot represents the two compared groups, either controls (C, in blue) or 22q11.2DS (del, in red). The lines represent smoothed methylation levels for each sample. Gene and CpG annotations are shown below the plot. We extended both up- and downstream of the DMR regions on the plot.

Noticeably, 9 DMPs (7 genes) were in the 22q11.2 region including 2 DMPs in *TBX1* (T-Box Transcription Factor 1). Both DMPs in *TBX1* were located on the promoter CpG island and hypomethylated in patients with 22q11.2 deletion. According to Wikipathway, the top gene enrichment is in the pathway associated with the 22q11.2 copy number variation syndrome (id: WP4657, q-value=0.009, 17 genes, **Table S3**). Based on GO, the gene enrichment analysis highlighted pathways related to cell fate, embryogenesis and various organ development, neuron migration (**Figure S2**, **Table S3**).

We found 15 genes consistent with the episignature reported by Rooney et al. [14] (enrichment p-value = 1.12e-6, marked in **Table S2**). The consistent genes included *FBXO41* (F-Box Protein 41) which shows 14 significant DMPs and was also detected in our DMRs analyses. This DMR (**Figure 3**), located on the second exon of *FBXO41*, was hypermethylated in 22q11.2DS patients compared to controls. Another gene, *SHANK2* (SH3 And Multiple Ankyrin Repeat Domains 2) was also reported by Rooney et al. Two DMPs in its promoter were hypomethylated in 22q11.2DS.

Then, we compared DNA methylation in the prefrontal cortex of mice with and without deletion (Tg vs. WT) and detected 165 DMPs located on 157 genes (**Figure S3**, **Table S5**). No DMR was identified (**Table S7**). GO enrichment pointed towards pathways related to myeloid cell differentiation, as well as regulation of synapse assembly (**Table S6**, **Figure S4**). Nine genes have been already identified by our DMPs and DMRs analyses in humans (**Table S5**).

### Differences in DNA methylation related to schizophrenia and stress

To explore the molecular pathways underlying SCZ in 22q11DS patients, we compared DNA methylation between nine patients with SCZ (age: 25.9 ± 8.1, F/M: 2/7) and five 22q11.2DS carriers without psychiatric disorders (age: 27.2 ± 12.2, F/M: 4/1) (**Table S1**). There was no difference between these two groups regarding age, sex nor ethnicity. We detected 3,957 DMPs on 3,240 genes (70.74% protein-coding genes, **Table S8, Figure 4**) and 15 significant DMRs on 10 genes (**Table S10**).

**Figure 4.**
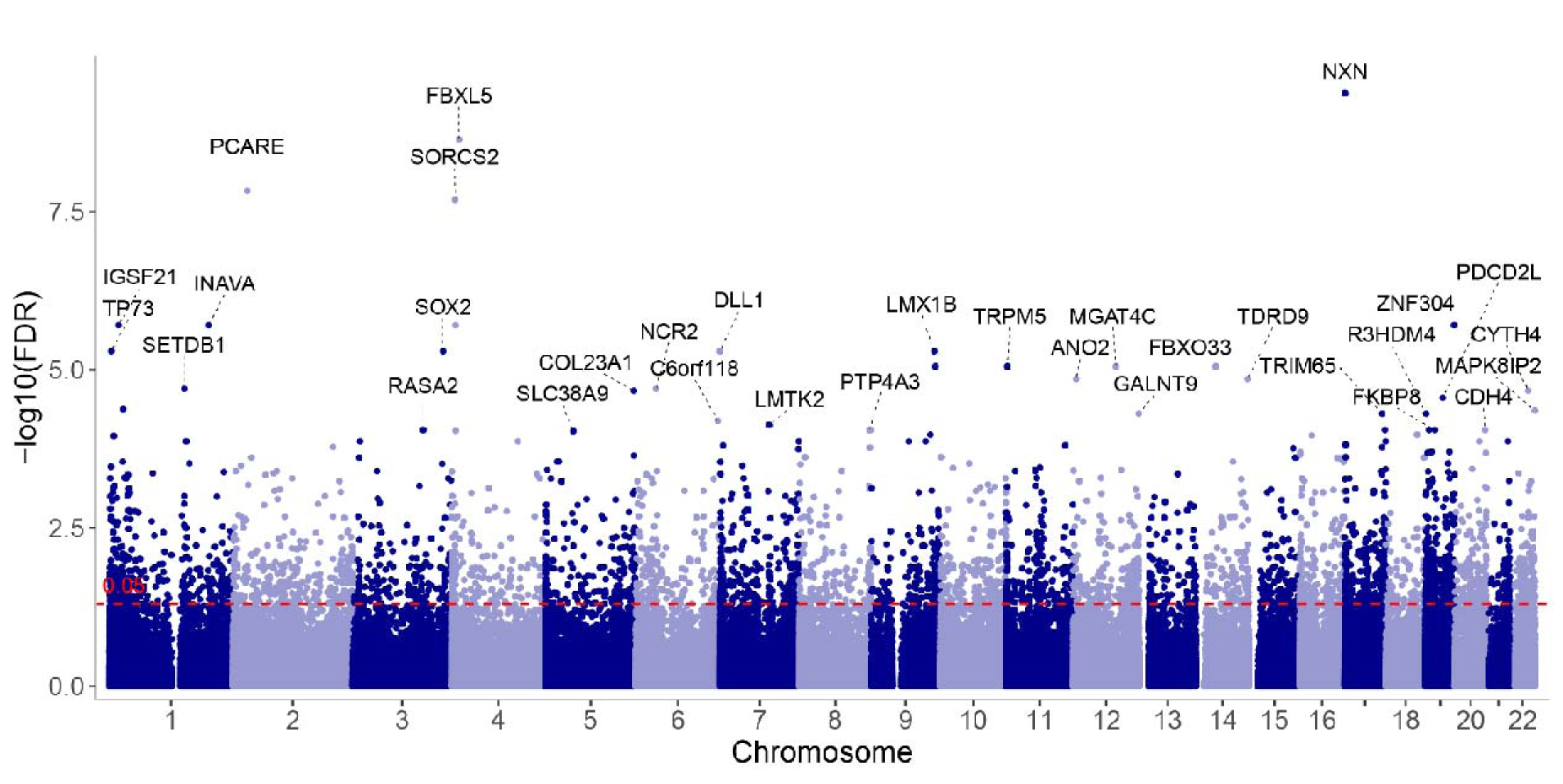
Manhattan plot of the differentially methylated CpGs identified from the comparison of individuals with 22q11DS with/without schizophrenia CpGs with FDR<1e-4 were labeled with their gene names.

We further explored the genes with significant DMPs (**Table S8)**: 356 genes overlapped with genes harboring DMPs related to 22q11.2DS (30.5% of the genes dysmethylated in the 22q11.2DS); 7 genes (*SEPTIN5*, *TBX1*, *DGCR6L*, *FAM230G*, *POM121L4P*, *PI4KA*, and *TMEM191C*) were located in the 22q11.2 region; 116 genes were reported by GWAS catalog as associated with SCZ representing a significant enrichment (q-value=1.31.10^-6^); MsigDB c3 predicted mir137-3p as the best regulator of them (97 genes, adjusted p-value=3.09.10^-6^); 127 genes were consistent with published SCZ EWAS studies in blood [57, 58] (enrichment p-value=4.72e-4); 4 genes have been already found in the EWAS analysis performed on neonatal dried blood spot samples of 22q11.2DS patients later suffering from psychiatric phenotypes [15]: *WDTC1, EFHD1*, *STK32C* and *PLEKHF1*. The genes associated to these DMPs are known to be highly expressed in the cerebral cortex, especially in neurons (p-value=2.57e-4), astrocytes (p-value=1.21e-3), and oligodendrocytes (p-value=1.21e-3). They were enriched in GO terms already identified when comparing individuals with 22q11.2DS and non-carriers, such as axonogenesis, embryonic organ development, and cell fate commitment (**Table S9**). However, new pathways emerged, including chemical synaptic transmission, channel activity, Wnt signaling, MAPK signaling, cognition, adult behavior (**Figure 5**).

**Figure 5.**
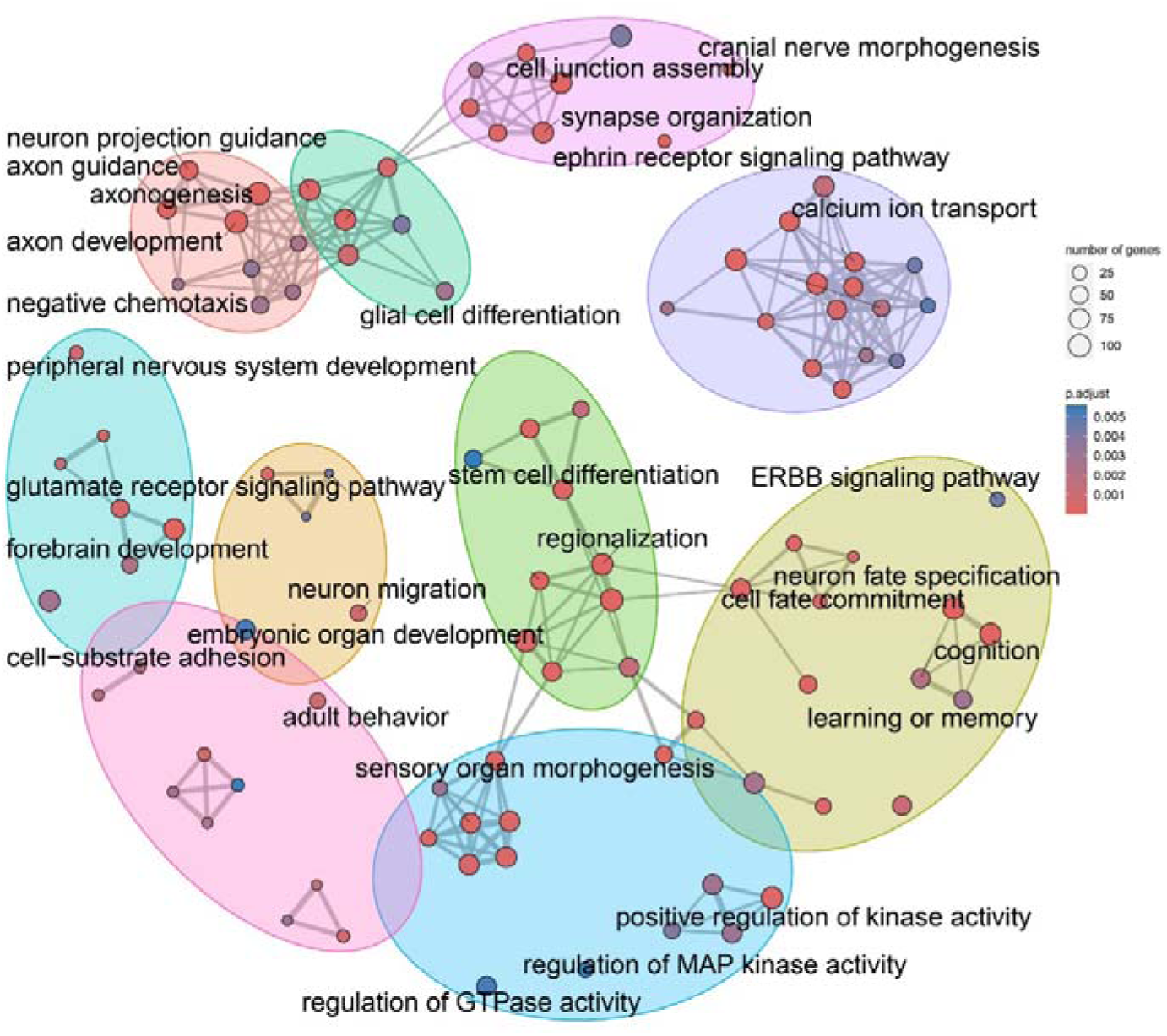
GO terms enrichment analysis from genes with differentially methylated CpGs in the comparison between individuals with 22q11.2DS and with/without SCZ. The color of the dots represents the adjusted p-value of the pathway and the size of the dots represents the number of genes with significant CpGs.The pathways were clustered into seven groups using the K-means method, and representative pathway names were retained

We directly compared the DNA methylation levels of the discordant twins (**Table S11a**). Genes with a CpG harboring a methylation difference > |50| were kept. Then, we retrieved genes with a difference in their expression measured by delta logCPM >|1.5| (**Table S11b**). The list contained 126 genes. According to WikiPathway, the enrichment is significant for the 22q11.2 copy number variation syndrome (id: WP4657; q-value=3.10^-15^; 16 genes, **Table S12**). When considering the unadjusted p-value, WikiPathway, KEGG and GO enrichment analyses pointed toward GABAergic synapse. The enrichment for the 22q11.2 copy number variation syndrome remained significant when selecting genes with methylation difference > |50| and delta logCPM >|2| (WikiPathway q-value=1.5.10^-24^).

As stress is a recognized environmental trigger for SCZ, we tested a double-hit model by comparing the prefrontal DNA methylation levels between transgenic mice (Df(h22q11)/+) exposed or not to an acute environmental stress. 416 DMPs located on 369 mouse genes and mapped to 275 human genes were detected (**Table S13**). These genes were enriched in GO terms similar to the ones identified when comparing 22q11.2DS with/without SCZ, including the Wnt signaling pathway (**Table S14**).

When overlapping the list of genes dysmethylated by stress in (Df(h22q11)/+) mice with the list of genes dysmethylated by SCZ in humans with 22q11.2DS, 62 genes were found in common. The pathway analysis on this overlap shows a significant enrichment for the Wnt receptor activity (GO:0042813; q-value =0.02). The enrichment with WikiPathway identified again a significant enrichment in the pathway 22q11 Copy Number Variation Syndrome (WP4657; q-value = 0.037; 4 genes involved: *TBX1*, *SEPTIN5*, *PI4KA*, *DGCR6L*).

### miRNA changes in 22q11.2 deletion carriers and in the context of SCZ

We also investigated miRNA expression differences between 22q11.2DS and non-carriers individuals as well as differences between 22q11.2DS patients with or without SCZ.

First, we compared individuals with 22q11.2DS and non carriers individuals. We did not detect significant differentially expressed miRNA after correction for multiple testing. However, eight miRNAs were hypoexpressed in 22q11.2DS with a nominal p-value<0.01 (FDR =0.518) (**Figure S5, Table S15**). When exploring these 8 miRNAs, we identified 1,942 unique target genes (**Table S16**). These target genes were enriched in pathways related to axonogenesis, synapse organization, and brain development, including some pathways that had been already identified by the DNA methylation comparison (**Table S17, Figure S6**). But more mechanistic pathways also emerged including BDNF signaling pathway, and response to glucocorticoids. Additionally, we observed that these target genes were enriched in genes identified in GWAS SCZ (q-value= 2.65e-04).

Second, we compared 22q11.2DS patients with or without SCZ. One miRNA was significantly hyperexpressed in 22q11.2DS patients with SCZ compared to those without SCZ (hsa-miR-6750-5p; q-value=1.24.10^-6^; log2FC=24.3) (**Figure S5**). Seven miRNAs were detected with nominal p-values < 0.01 (**Table S18**). These miRNAs have been associated with 1,256 unique target genes (**Table S19**). Enrichment analysis showed that these genes were associated with neurotransmitter secretion, synapse organization but also Wnt signaling pathway (**Figure S7, Table S20**). These genes are known to be highly expressed in the cerebral cortex (FDR=8.70e-6).

### Overlap between DNA methylation and miRNA expression analyses

Among the genes targeted by the miRNA differentially expressed in individuals with 22q11.2DS compared to non-carriers, 102 were also dysmethylated when comparing carriers and non-carriers. Thus, these genes are dysregulated by both epigenetic mechanisms in the context of the 22q11.2 deletion. They are nominally enriched in the regulation of neuron projection development (9 genes represented, q-value=0.3) (**Table S21**). The overlap of genes identified in the context of the deletion by DNA methylation in humans, DNA methylation in mice, and miRNA expression in humans highlighted only one gene, namely *NTN1* (**Figure 6**). This gene is important in maintaining excitatory connectivity in the adult mesolimbic system [59].

**Figure 6:**
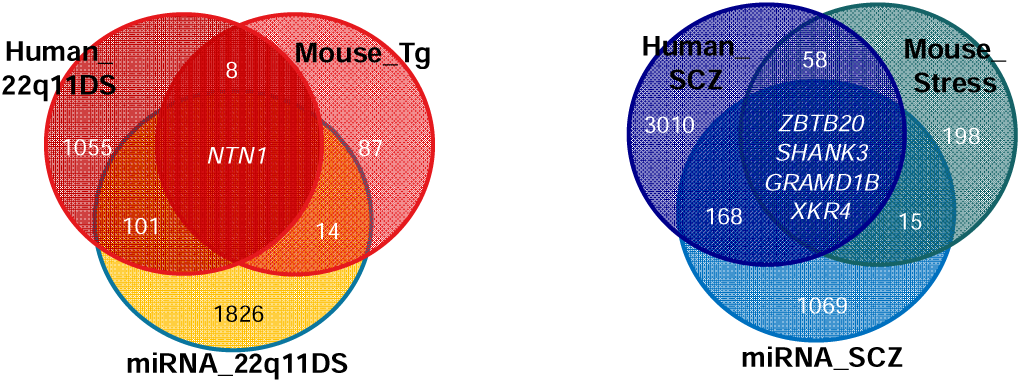
Venn plot of genes identified in the different analyses. The left diagram shows the overlap between genes with differentially methylated CpGs in individuals with 22q11DS compared to non-carriers (human_22q11DS), genes targeted by miRNA differentially expressed in individuals with 22q11DS compared to non-carriers (miRNA_22q11DS), and genes with differentially methylated CpGs in mice with the deletion compared to mice without the deletion (Mouse_Tg). The right diagram shows the overlap between genes with differentially methylated CpGs in individuals with 22q11DS when comparing those with and without SCZ (human_SCZ), genes targeted by miRNA differentially expressed in individuals with 22q11DS when comparing those with and without SCZ (miRNA_SCZ), and genes with differentially methylated CpGs in mice with the deletion when comparing those exposed or not to stress (Mouse_Stress).

Among the genes targeted by the miRNA differentially expressed in individuals 22q11.2DS with/without SCZ, 172 were also dysmethylated in the same comparison. Thus, these genes are dysregulated by both epigenetic mechanisms in the context of SCZ in 22q11.2DS. Among the enriched pathways detected from these genes, Wnt signaling is one of the strongest (54 genes represented, q-value=0.001) (**Table S22)**. The overlap of genes identified by DNA methylation in the context of SCZ in humans, in the context of stress in mice, and by miRNA expression in humans highlighted four genes (**Figure 6**), three of them being already associated with SCZ according to GWAS catalog (*ZBTB20, SHANK3, GRAMD1B*). *XKR4* has been previously associated with the response to one antipsychotic treatment in schizophrenia [60].

## Discussion

Through this translational study, we aimed to identify how the epigenetic changes induced by the 22q11.2 deletion could lead to SCZ. First, we investigated the effects of the deletion on the DNA methylation in humans and transgenic mice. Some genes in the 22q11.2 region were dysmethylated including *TBX1* (T-Box Transcription Factor 1). Two CpGs from its promoter were hypomethylated in human carriers compared to non-carriers. *TBX1* point mutations have been reported to result in most of the physical features of the 22q11.2 deletion syndrome phenotype in humans [8]. But *TBX1* is a transcription factor regulating more than 2,000 genes [7]. This is consistent with the diffuse impact of the deletion on DNA methylation with hits identified in every human and mouse chromosomes. This result fuels the hypothesis of 22q11.2DS being a primary transcription deregulation syndrome [61]. In our case, the dysmethylated genes belong to pathways related to broad development, some of them being associated with extra-psychiatric phenotype (i.e. cardiac defect, blood cell differentiation), in accordance with the multi-systemic consequences of the syndrome. However, neuron migration or axonogenesis were also included confirming the impact of the deletion on nervous system development. Interestingly, some of our findings are concordant with previous comparisons between 22q11.2DS patients and non-carriers. Indeed, we found an hypermethylation in the body of *FBXO41*, and an hypomethylation in the promoter of *SHANK2*, as reported in a previously published episignature [14]. *FBXO41* is highly expressed in the brain, especially in neurons [62]. It has been identified as a GWAS signal of educational attainment and intelligence, and it has a diminished expression in SCZ patients according to PsychENCODE. *SHANK2* is a postsynaptic scaffolding protein of excitatory synapses associated with susceptibility to neurodevelopmental disorders [63], SCZ [64], bipolar disorder [65], and other neuropsychiatric disorders [66]. Dysmethylation of these genes could play a role in the risk of developing psychiatric disorders.

Moreover, to examine how SCZ is associated with 22q11.2DS, we conducted a comparison of DNA methylation differences between individuals with and without SCZ in the context of 22q11.2DS. Some of our results confirmed previous findings. One region located in the intron of the gene *PTPRN2* (Protein Tyrosine Phosphatase Receptor Type N2) was hypermethylated in 22q11.2DS patients with SCZ. In a previous Methylation-Wide Association Study (MWAS) study comparing 22q11.2DS with/without SCZ, this gene has been already highlighted [13]. It is believed to play a role in organizing the cytomatrix at the active zone of nerve terminals, thereby regulating neurotransmitter release, and has been identified as a gene of interest in case-case GWAS of SCZ and bipolar disorder [67]. Four of the genes dysmethylated in the present study have been documented as dysmethylated prior to the onset of SCZ in individuals with 22q11 at birth, implying that certain methylation patterns may precede the manifestation of symptoms. A notable portion of the identified genes has previously been linked to SCZ through GWAS or MWAS. Additionally, many of these genes were regulated by mir137-3p, a member of mir137 family, a significant miRNA associated with SCZ [68]. These results suggest that the development of SCZ in individuals with del22q11.2 may share common underlying pathological mechanisms with SCZ in the general population.

The newly identified pathways related to SCZ in 22q11.2DS partly overlap those detected by the sole effects of 22q11.2DS. However, novel pathways have emerged, encompassing chemical synaptic transmission, channel activity, Wnt signaling, and MAPK signaling. To explore the potential role of stress in these new dysregulations, we investigated its impact using a gene x environment interaction mouse model. Stress induced DNA methylation alterations in the prefrontal cortex of Df(h22q11)/+ mice, particularly in genes associated with 22q11.2DS and the Wnt pathways. These findings suggest that the onset of SCZ may involve not only an exacerbation of pathways previously affected by the deletion but also new DNA methylation changes linked to environmental stressors.

Finally, given that certain crucial miRNAs associated with SCZ are situated within the 22q11.2 region, and considering that DGCR8, the gene encoding a protein essential for miRNA processing, is also situated in this region, we conducted an analysis of miRNA expression in individuals with and without SCZ within the context of 22q11.2DS. Probably due to limited power, only one significant result was identified: hsa-miR-6750-5p is over-expressed in 22q11.2 carriers with SCZ compared to carriers without SCZ. By relaxing the level of significance and imputing the gene targets, we found an overlap with dysmethylated genes. Again, genes involved in brain development were associated with the effects of the deletion whereas the effects of SCZ highlighted genes from the Wnt pathway.

Four genes were at the overlap of the three comparisons (DNA methylation in individuals with 22q11.2DS with/without SCZ, miRNA expression in individuals with 22q11.2DS with/without SCZ, effect of stress on DNA methylation in Tg mice). These genes are very good candidates to explain the emergence of SCZ in the context of 22q11.2DS through the effect of stress. Three of them have already been associated with SCZ. *ZBTB20* (Zinc Finger and BTB Domain Containing 20) acts as a transcriptional repressor and plays a role in neurogenesis by modulating the sequential generation of neuronal layers in the developing cortex [69]. *SHANK3* (SH3 And Multiple Ankyrin Repeat Domains 3) encodes multidomain scaffold proteins of the postsynaptic density and plays a role in synapse formation and dendritic spine maturation. It has been implicated in the mechanisms underlying autism spectrum disorder and SCZ [70, 71]. *GRAMD1B* (GRAM Domain Containing 1B) is involved in the cellular response to cholesterol and is highly expressed in the brain. *XKR4* (XK Related 4) is also strongly expressed in the brain, especially in neurons and oligodendrocyte precursor cells. It can expose phosphatidylserine as an “eat me” signal to phagocytes and removed unwanted synapses [72].

Our study has certain limitations. Primarily, due to to the restricted sample size, it is imperative to interpret the results with caution. Unexpected but explainable, some of the published differentially expressed miRNAs in SCZ (e.g., mir137) were not expressed in our samples, maybe resulting from the deletion of DGCR8. However, some findings align with existing literature and indicate pathways related to 22q11.2DS, bolstering confidence in the analyses. Utilizing a genotype-first cohort could mitigate heterogeneity and enhance the likelihood of detecting true positives. Notably, we did not investigate the precise boundaries of the 22q11 deletions, introducing a potential bias as various types of deletions exist. Nonetheless, there is no clear correlation between deletion size and the SCZ phenotype [73], and it is highly likely that the majority of subjects carried the typically deleted 3Mb region, which accounts for 90% of cases in clinical practice. The inclusion of a pair of discordant monozygotic twins enhances the robustness of the comparison. Additionally, our human comparisons are constrained by the peripheral approach, as DNA methylation in the blood may not perfectly reflect patterns in the brain. However, validating our findings with data from brain tissues in an animal model provides a powerful supplement.

This translational study has investigated the epigenetic factors related to SCZ in the context of 22q11.2DS. It highlighted the potential interest of the Wnt pathway, mediated by environmental stress, to explain the incomplete penetrance of SCZ in this genetic condition. Disentangling the molecular mechanisms related to the gene x environment interactions in psychiatry could be highly beneficial for clinicians and patients. The emergence of SCZ in 22q11.2DS is challenging for psychiatrists [74, 75] and identification of biomarkers for early diagnosis among del22q11.2 carriers as well as exploration of novel therapeutic targets would be highly useful.

## Data Availability

All data produced in the present study are available upon reasonable request to the authors

## Acknowledgements

We would like to thank all the patients and their families. This work was funded by ANR Epi-Young (ANR-17-CE37-0003-01) and Fédération pour la Recherche sur le Cerveau. It has been supported by the French government’s “Investissements d’Avenir” programme (ANR-18-RHUS-0014 PsyCARE), French Ministry grants PHRC AOM07-118 (ICAAR) and PHRC 13-0681 (START). Boris Chaumette received a grant from Fondation Bettencourt Schueller (CCA INSERM Bettencourt). TMJ received the Df(h22q11)/+ mice as a part of the Innovative Medicines Initiative Joint Undertaking (IMI) under Grant Agreement No. 115008. Additional support for AT came from grants from Servier. FD received support from Fondation pour la Recherche Médicale (FRM). We acknowledge the use of the Biological Resource Center NSPN, GHU Paris Psychiatrie & Neurosciences Biobank (BB-0033-00026).

## Data Availability Statement

The data supporting this study and the script containing each analysis are available upon reasonable request.

## References

1. Marshall CR, Howrigan DP, Merico D, Thiruvahindrapuram B, Wu W, Greer DS, et al. Contribution of copy number variants to schizophrenia from a genome-wide study of 41,321 subjects. Nat Genet. 2017;49:27–35.

2. Cortés-Martín J, Peñuela NL, Sánchez-García JC, Montiel-Troya M, Díaz-Rodríguez L, Rodríguez-Blanque R. Deletion Syndrome 22q11.2: A Systematic Review. Children (Basel). 2022;9:1168.

3. Olsen L, Sparsø T, Weinsheimer SM, Santos MBQD, Mazin W, Rosengren A, et al. Prevalence of rearrangements in the 22q11.2 region and population-based risk of neuropsychiatric and developmental disorders in a Danish population: a case-cohort study. The Lancet Psychiatry. 2018;5:573–580.

4. Schneider M, Debbané M, Bassett AS, Chow EWC, Fung WLA, van den Bree M, et al. Psychiatric disorders from childhood to adulthood in 22q11.2 deletion syndrome: results from the International Consortium on Brain and Behavior in 22q11.2 Deletion Syndrome. Am J Psychiatry. 2014;171:627–639.

5. Bassett AS. Clinical genetics of schizophrenia and related neuropsychiatric disorders. Psychiatry Res. 2023;319:114992.

6. McDonald-McGinn DM, Sullivan KE, Marino B, Philip N, Swillen A, Vorstman JAS, et al. 22q11.2 deletion syndrome. Nat Rev Dis Primers. 2015;1:15071.

7. Du Q, de la Morena MT, van Oers NSC. The Genetics and Epigenetics of 22q11.2 Deletion Syndrome. Frontiers in Genetics. 2020;10:1365.

8. Zinkstok JR, Boot E, Bassett AS, Hiroi N, Butcher NJ, Vingerhoets C, et al. Neurobiological perspective of 22q11.2 deletion syndrome. The Lancet Psychiatry. 2019;6:951–960.

9. Bassett AS, Lowther C, Merico D, Costain G, Chow EWC, van Amelsvoort T, et al. Rare Genome-Wide Copy Number Variation and Expression of Schizophrenia in 22q11.2 Deletion Syndrome. Am J Psychiatry. 2017:appiajp201716121417.

10. Cleynen I, Engchuan W, Hestand MS, Heung T, Holleman AM, Johnston HR, et al. Genetic contributors to risk of schizophrenia in the presence of a 22q11.2 deletion. Mol Psychiatry. 2021;26:4496–4510.

11. Alver M, Mancini V, Läll K, Schneider M, Romano L, Estonian Biobank Research Team, et al. Contribution of schizophrenia polygenic burden to longitudinal phenotypic variance in 22q11.2 deletion syndrome. Mol Psychiatry. 2022;27:4191–4200.

12. Singh SM, Murphy B, O’Reilly R. Monozygotic twins with chromosome 22q11 deletion and discordant phenotypes: updates with an epigenetic hypothesis. J Med Genet. 2002;39:e71.

13. Carmel M, Michaelovsky E, Weinberger R, Frisch A, Mekori-Domachevsky E, Gothelf D, et al. Differential methylation of imprinting genes and MHC locus in 22q11.2 deletion syndrome-related schizophrenia spectrum disorders. World J Biol Psychiatry. 2021;22:46–57.

14. Rooney K, Levy MA, Haghshenas S, Kerkhof J, Rogaia D, Tedesco MG, et al. Identification of a DNA Methylation Episignature in the 22q11.2 Deletion Syndrome. International Journal of Molecular Sciences. 2021;22.

15. Starnawska A, Hansen CS, Sparsø T, Mazin W, Olsen L, Bertalan M, et al. Differential DNA methylation at birth associated with mental disorder in individuals with 22q11.2 deletion syndrome. Transl Psychiatry. 2017;7:e1221.

16. Kebir O, Chaumette B, Rivollier F, Miozzo F, Lemieux Perreault LP, Barhdadi A, et al. Methylomic changes during conversion to psychosis. Mol Psychiatry. 2017;22:512–518.

17. Iftimovici A, He Q, Jiao C, Duchesnay E, Krebs M-O, Kebir O, et al. Longitudinal MicroRNA Signature of Conversion to Psychosis. Schizophr Bull. 2024;50:363–373.

18. Shang R, Lee S, Senavirathne G, Lai EC. microRNAs in action: biogenesis, function and regulation. Nat Rev Genet. 2023:1–18.

19. Cui Y, Lyu X, Ding L, Ke L, Yang D, Pirouz M, et al. Global miRNA dosage control of embryonic germ layer specification. Nature. 2021;593:602–606.

20. Xu B, Hsu P-K, Stark KL, Karayiorgou M, Gogos JA. Derepression of a Neuronal Inhibitor due to miRNA Dysregulation in a Schizophrenia-Related Microdeletion. Cell. 2013;152:262–275.

21. Stark KL, Xu B, Bagchi A, Lai W-S, Liu H, Hsu R, et al. Altered brain microRNA biogenesis contributes to phenotypic deficits in a 22q11-deletion mouse model. Nat Genet. 2008;40:751– 760.

22. Earls LR, Fricke RG, Yu J, Berry RB, Baldwin LT, Zakharenko SS. Age-Dependent MicroRNA Control of Synaptic Plasticity in 22q11 Deletion Syndrome and Schizophrenia. J Neurosci. 2012;32:14132–14144.

23. Ying S, Heung T, Zhang Z, Yuen RKC, Bassett AS. Schizophrenia Risk Mediated by microRNA Target Genes Overlapped by Genome-Wide Rare Copy Number Variation in 22q11.2 Deletion Syndrome. Front Genet. 2022;13:812183.

24. de la Morena MT, Eitson JL, Dozmorov IM, Belkaya S, Hoover AR, Anguiano E, et al. Signature MicroRNA expression patterns identified in humans with 22q11.2 deletion/DiGeorge syndrome. Clin Immunol. 2013;147:11–22.

25. Sellier C, Hwang VJ, Dandekar R, Durbin-Johnson B, Charlet-Berguerand N, Ander BP, et al. Decreased DGCR8 Expression and miRNA Dysregulation in Individuals with 22q11.2 Deletion Syndrome. PLoS ONE. 2014;9:e103884.

26. Thomas KT, Zakharenko SS. MicroRNAs in the Onset of Schizophrenia. Cells. 2021;10:2679.

27. Rey R, Suaud-Chagny M-F, Dorey J-M, Teyssier J-R, d’Amato T. Widespread transcriptional disruption of the microRNA biogenesis machinery in brain and peripheral tissues of individuals with schizophrenia. Transl Psychiatry. 2020;10:1–13.

28. Cillo F, Coppola E, Habetswallner F, Cecere F, Pignata L, Toriello E, et al. Understanding the Variability of 22q11.2 Deletion Syndrome: The Role of Epigenetic Factors. Genes. 2024;15:321.

29. Sabaie H, Gharesouran J, Asadi MR, Farhang S, Ahangar NK, Brand S, et al. Downregulation of miR-185 is a common pathogenic event in 22q11.2 deletion syndrome-related and idiopathic schizophrenia. Metab Brain Dis. 2022;37:1175–1184.

30. Ilen L, Feller C, Eliez S, Schneider M. Increased affective reactivity to daily social stressors is associated with more severe psychotic symptoms in youths with 22q11.2 deletion syndrome. Psychol Med. 2023;53:1–12.

31. Zannas AS, Chrousos GP. Epigenetic programming by stress and glucocorticoids along the human lifespan. Mol Psychiatry. 2017;22:640–646.

32. Chaumette B, Kebir O, Mam-Lam-Fook C, Morvan Y, Bourgin J, Godsil BP, et al. Salivary cortisol in early psychosis: New findings and meta-analysis. Psychoneuroendocrinology. 2016;63:262– 270.

33. Cullen AE, Labad J, Oliver D, Al-Diwani A, Minichino A, Fusar-Poli P. The Translational Future of Stress Neurobiology and Psychosis Vulnerability: A Review of the Evidence. Curr Neuropharmacol. 2024;22:350–377.

34. Jonas RK, Montojo CA, Bearden CE. The 22q11.2 Deletion Syndrome as a Window into Complex Neuropsychiatric Disorders Over the Lifespan. Biological Psychiatry. 2014;75:351–360.

35. Karayiorgou M, Simon TJ, Gogos JA. 22q11.2 microdeletions: linking DNA structural variation to brain dysfunction and schizophrenia. Nature Reviews Neuroscience. 2010;11:402–416.

36. Meechan DW, Maynard TM, Tucker ES, Fernandez A, Karpinski BA, Rothblat LA, et al. Modeling a model: Mouse genetics, 22q11.2 Deletion Syndrome, and disorders of cortical circuit development. Progress in Neurobiology. 2015;130:1–28.

37. Tripathi A, Spedding M, Schenker E, Didriksen M, Cressant A, Jay TM. Cognition- and circuit-based dysfunction in a mouse model of 22q11.2 microdeletion syndrome: effects of stress. Translational Psychiatry. 2020;10:1–15.

38. Tripathi A, Schenker E, Spedding M, Jay TM. The hippocampal to prefrontal cortex circuit in mice: a promising electrophysiological signature in models for psychiatric disorders. Brain Structure and Function. 2016;221:2385–2391.

39. Meissner A, Gnirke A, Bell GW, Ramsahoye B, Lander ES, Jaenisch R. Reduced representation bisulfite sequencing for comparative high-resolution DNA methylation analysis. Nucleic Acids Res. 2005;33:5868–5877.

40. Krueger F, Andrews SR. Bismark: a flexible aligner and methylation caller for Bisulfite-Seq applications. Bioinformatics. 2011;27:1571–1572.

41. Hicks SC, Irizarry RA. methylCC: technology-independent estimation of cell type composition using differentially methylated regions. Genome Biology. 2019;20:261.

42. Feng H, Wu H. Differential methylation analysis for bisulfite sequencing using DSS. Quant Biol. 2019;7:327–334.

43. Ritchie ME, Phipson B, Wu D, Hu Y, Law CW, Shi W, et al. limma powers differential expression analyses for RNA-sequencing and microarray studies. Nucleic Acids Research. 2015;43:e47–e47.

44. Peters TJ, Buckley MJ, Chen Y, Smyth GK, Goodnow CC, Clark SJ. Calling differentially methylated regions from whole genome bisulphite sequencing with DMRcate. Nucleic Acids Res. 2021;49:e109.

45. Leek JT, Johnson WE, Parker HS, Jaffe AE, Storey JD. The sva package for removing batch effects and other unwanted variation in high-throughput experiments. Bioinformatics. 2012;28:882–883.

46. Chen Y, McCarthy D, Baldoni P, Robinson M, Smyth G. edgeR: differential analysis of sequence read count data User’s Guide.

47. Bolger AM, Lohse M, Usadel B. Trimmomatic: a flexible trimmer for Illumina sequence data. Bioinformatics. 2014;30:2114–2120.

48. Dobin A, Davis CA, Schlesinger F, Drenkow J, Zaleski C, Jha S, et al. STAR: ultrafast universal RNA-seq aligner. Bioinformatics. 2013;29:15–21.

49. Li B, Dewey CN. RSEM: accurate transcript quantification from RNA-Seq data with or without a reference genome. BMC Bioinformatics. 2011;12:323.

50. Love MI, Huber W, Anders S. Moderated estimation of fold change and dispersion for RNA-seq data with DESeq2. Genome Biology. 2014;15:550.

51. Chen Y, Wang X. miRDB: an online database for prediction of functional microRNA targets. Nucleic Acids Res. 2020;48:D127–D131.

52. Chen Y, Wang X. miRDB: an online database for prediction of functional microRNA targets. Nucleic Acids Research. 2020;48:D127–D131.

53. clusterProfiler: an R Package for Comparing Biological Themes Among Gene Clusters | OMICS: A Journal of Integrative Biology. https://www.liebertpub.com/doi/10.1089/omi.2011.0118. Accessed 19 September 2023.

54. Leeuw CA de, Mooij JM, Heskes T, Posthuma D. MAGMA: Generalized Gene-Set Analysis of GWAS Data. PLOS Computational Biology. 2015;11:e1004219.

55. Watanabe K, Taskesen E, van Bochoven A, Posthuma D. Functional mapping and annotation of genetic associations with FUMA. Nat Commun. 2017;8:1826.

56. Jain A, Tuteja G. TissueEnrich: Tissue-specific gene enrichment analysis. Bioinformatics. 2019;35:1966–1967.

57. Hannon E, Dempster E, Viana J, Burrage J, Smith AR, Macdonald R, et al. An integrated genetic-epigenetic analysis of schizophrenia: evidence for co-localization of genetic associations and differential DNA methylation. Genome Biology. 2016;17.

58. Montano C, Taub MA, Jaffe A, Briem E, Feinberg JI, Trygvadottir R, et al. Association of DNA Methylation Differences With Schizophrenia in an Epigenome-Wide Association Study. JAMA Psychiatry. 2016;73:506–514.

59. Cline MM, Juarez B, Hunker A, Regiarto EG, Hariadi B, Soden ME, et al. Netrin-1 regulates the balance of synaptic glutamate signaling in the adult ventral tegmental area. eLife;12:e83760.

60. Lavedan C, Licamele L, Volpi S, Hamilton J, Heaton C, Mack K, et al. Association of the NPAS3 gene and five other loci with response to the antipsychotic iloperidone identified in a whole genome association study. Mol Psychiatry. 2009;14:804–819.

61. Meechan DW, Maynard TM, Gopalakrishna D, Wu Y, LaMantia A-S. When Half Is Not Enough: Gene Expression and Dosage in the 22q11 Deletion Syndrome. Gene Expr. 2018;13:299–310.

62. hg38 Single cell RNA expression levels cell types from many organs (FBXO41). https://genome.ucsc.edu/cgi-bin/hgc?g=singleCellMerged&i=FBXO41&hgsid=1770785272_5mWVkcUdsAmLTNxTdhvh1mDdDK3Q&singleCellMerged_facet_op=add&singleCellMerged_facet_fieldName=stage&singleCellMerged_facet_fieldVal=fetal&singleCellMerged_page=1#singleCellMerged_self_a_stage. Accessed 14 November 2023.

63. Leblond CS, Nava C, Polge A, Gauthier J, Huguet G, Lumbroso S, et al. Meta-analysis of SHANK Mutations in Autism Spectrum Disorders: A Gradient of Severity in Cognitive Impairments. PLoS Genet. 2014;10:e1004580.

64. Peykov S, Berkel S, Schoen M, Weiss K, Degenhardt F, Strohmaier J, et al. Identification and functional characterization of rare SHANK2 variants in schizophrenia. Mol Psychiatry. 2015;20:1489–1498.

65. Stahl EA, Breen G, Forstner AJ, McQuillin A, Ripke S, Trubetskoy V, et al. Genome-wide association study identifies 30 Loci Associated with Bipolar Disorder. Nat Genet. 2019;51:793– 803.

66. Homann OR, Misura K, Lamas E, Sandrock RW, Nelson P, McDonough SI, et al. Whole-genome sequencing in multiplex families with psychoses reveals mutations in the SHANK2 and SMARCA1 genes segregating with illness. Mol Psychiatry. 2016;21:1690–1695.

67. Curtis D, Vine A, McQuillin A, Bass N, Pereira A, Kandaswamy R, et al. Case-case genome wide association analysis reveals markers differentially associated with schizophrenia and bipolar disorder and implicates calcium channel genes. Psychiatr Genet. 2011;21:1–4.

68. Sakamoto K, Crowley JJ. A comprehensive review of the genetic and biological evidence supports a role for MicroRNA-137 in the etiology of schizophrenia. Am J Med Genet B Neuropsychiatr Genet. 2018;177:242–256.

69. Tonchev AB, Tuoc TC, Rosenthal EH, Studer M, Stoykova A. Zbtb20 modulates the sequential generation of neuronal layers in developing cortex. Mol Brain. 2016;9:65.

70. Zhou Y, Kaiser T, Monteiro P, Zhang X, Van der Goes MarieS, Wang D, et al. Mice with Shank3 Mutations Associated with ASD and Schizophrenia Display Both Shared and Distinct Defects. Neuron. 2016;89:147–162.

71. Gauthier J, Champagne N, Lafrenière RG, Xiong L, Spiegelman D, Brustein E, et al. De novo mutations in the gene encoding the synaptic scaffolding protein SHANK3 in patients ascertained for schizophrenia. Proceedings of the National Academy of Sciences. 2010;107:7863–7868.

72. Maruoka M, Zhang P, Mori H, Imanishi E, Packwood DM, Harada H, et al. Caspase cleavage releases a nuclear protein fragment that stimulates phospholipid scrambling at the plasma membrane. Molecular Cell. 2021;81:1397–1410.e9.

73. Michaelovsky E, Frisch A, Carmel M, Patya M, Zarchi O, Green T, et al. Genotype-phenotype correlation in 22q11.2 deletion syndrome. BMC Med Genet. 2012;13:122.

74. Iftimovici A, Krebs M-O, Chaumette B. Clinical management of psychosis in 22q11.2 deletion syndrome. J Psychiatry Neurosci. 2022;47:E391–E392.

75. Boot E, Óskarsdóttir S, Loo JCY, Crowley TB, Orchanian-Cheff A, Andrade DM, et al. Updated clinical practice recommendations for managing adults with 22q11.2 deletion syndrome. Genet Med. 2023;25:100344.

