## Supplementary figures for "Epigenetic factors in the 22q11.2 deletion syndrome in relation to stress and schizophrenia"

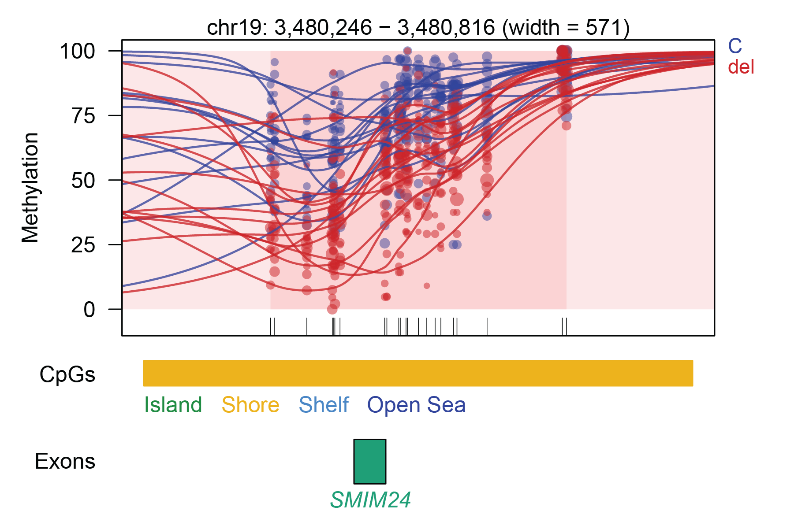

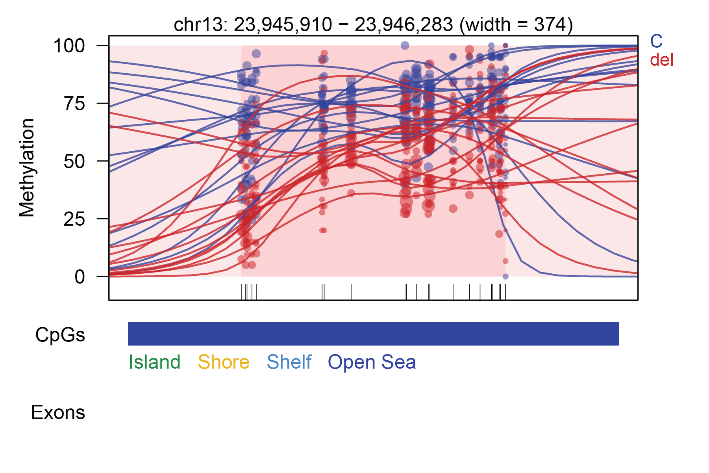

**Figure S1. 10 DMRs plots in 22q11.2 deletion compared to non-carriers**

Each dot represents the methylation level of an individual CpG in a single sample, where the size of the dot represents coverage, and the color of the dot represents the two compared groups, either controls (C, in blue) or 22q11.2DS (del, in red). The lines represent smoothed methylation levels for each sample. Gene and CpG annotations are shown below the plot. We extended both up- and downstream of the DMR regions on the plot.

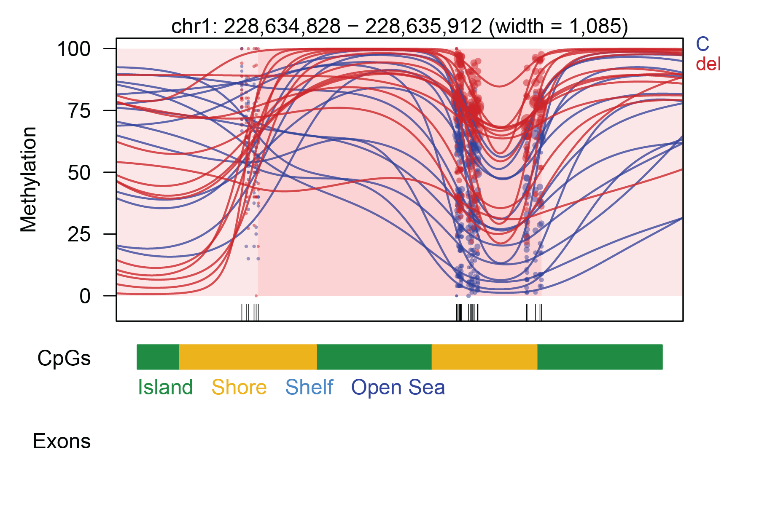

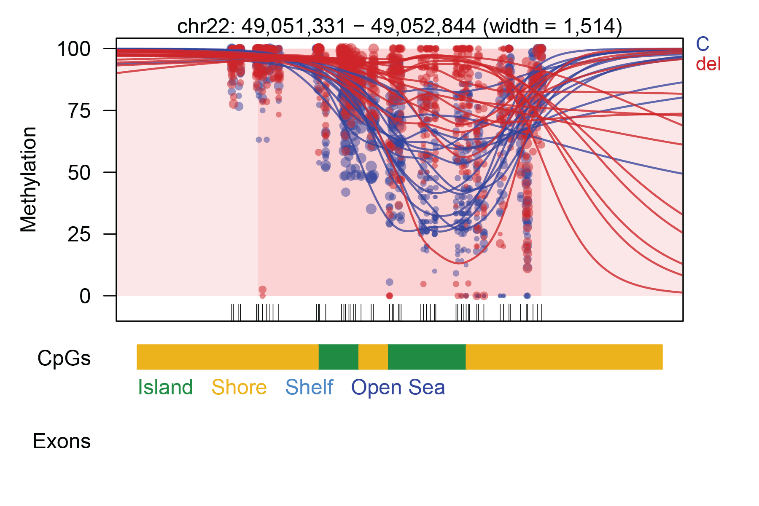

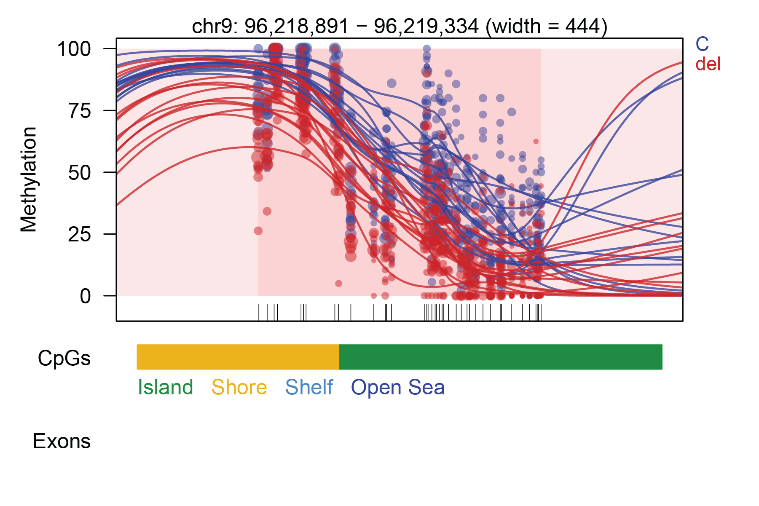

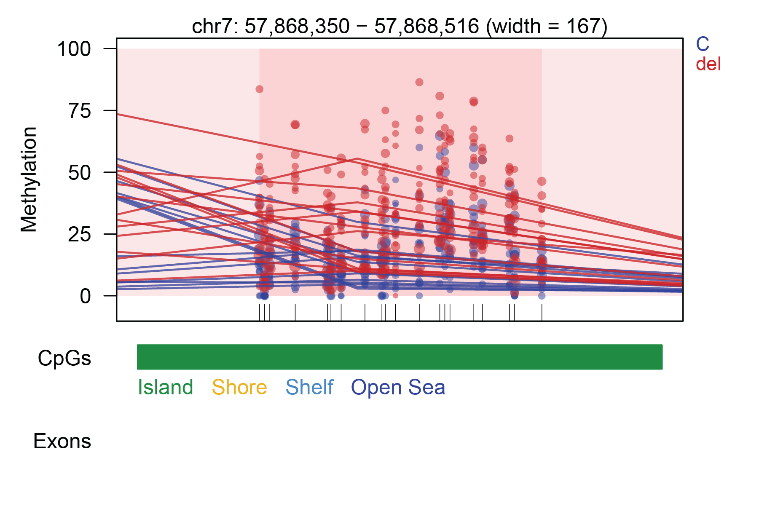

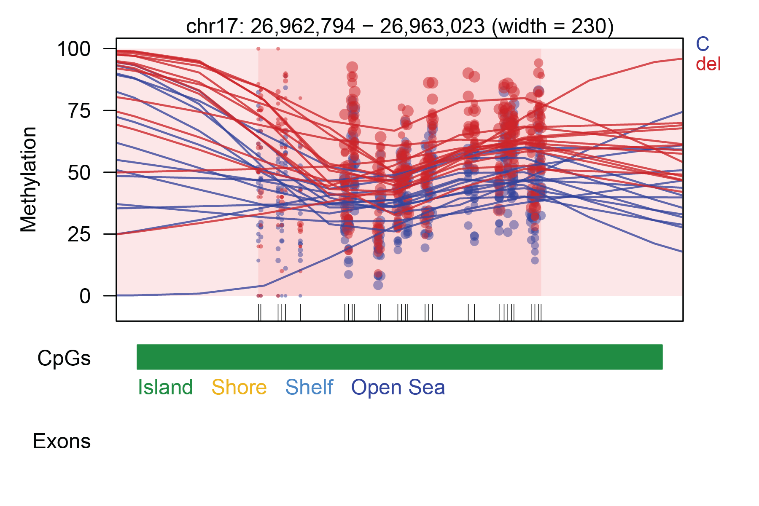

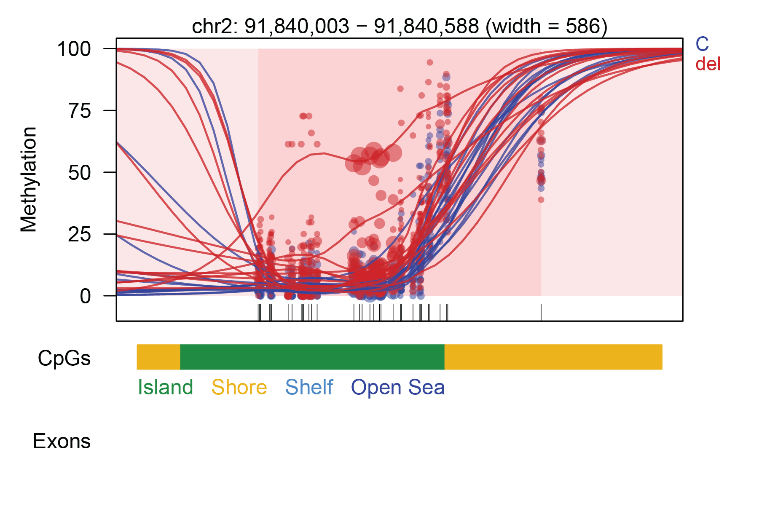

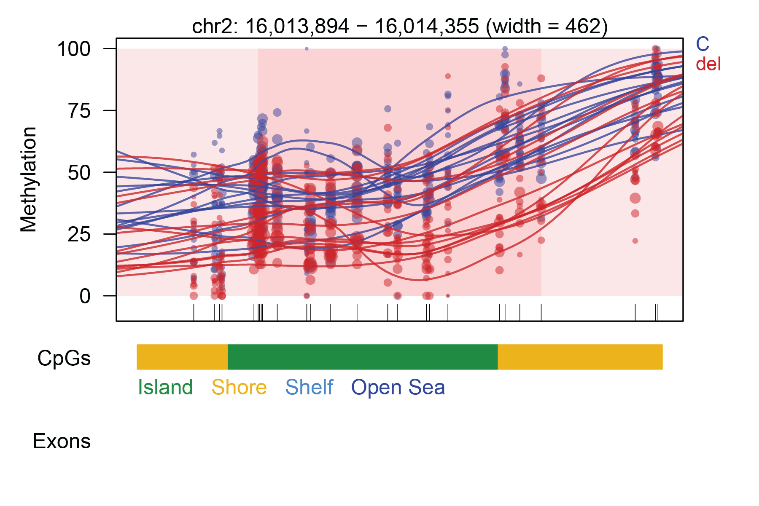

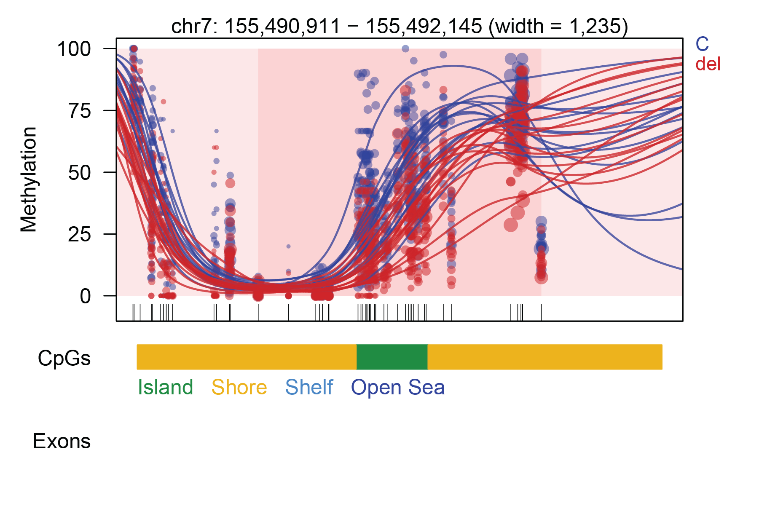
**Figure S2 Gene Ontology enrichment analysis from the Differentially Methylated Positions identified in the comparison between 22q11.2DS individuals and non-carriers**

The color of the dots represents the adjusted p-value of the pathway and the size of the dots represents the number of genes in each pathway. The pathways were clustered into seven groups using the K-means method, and representative pathway names were labeled.

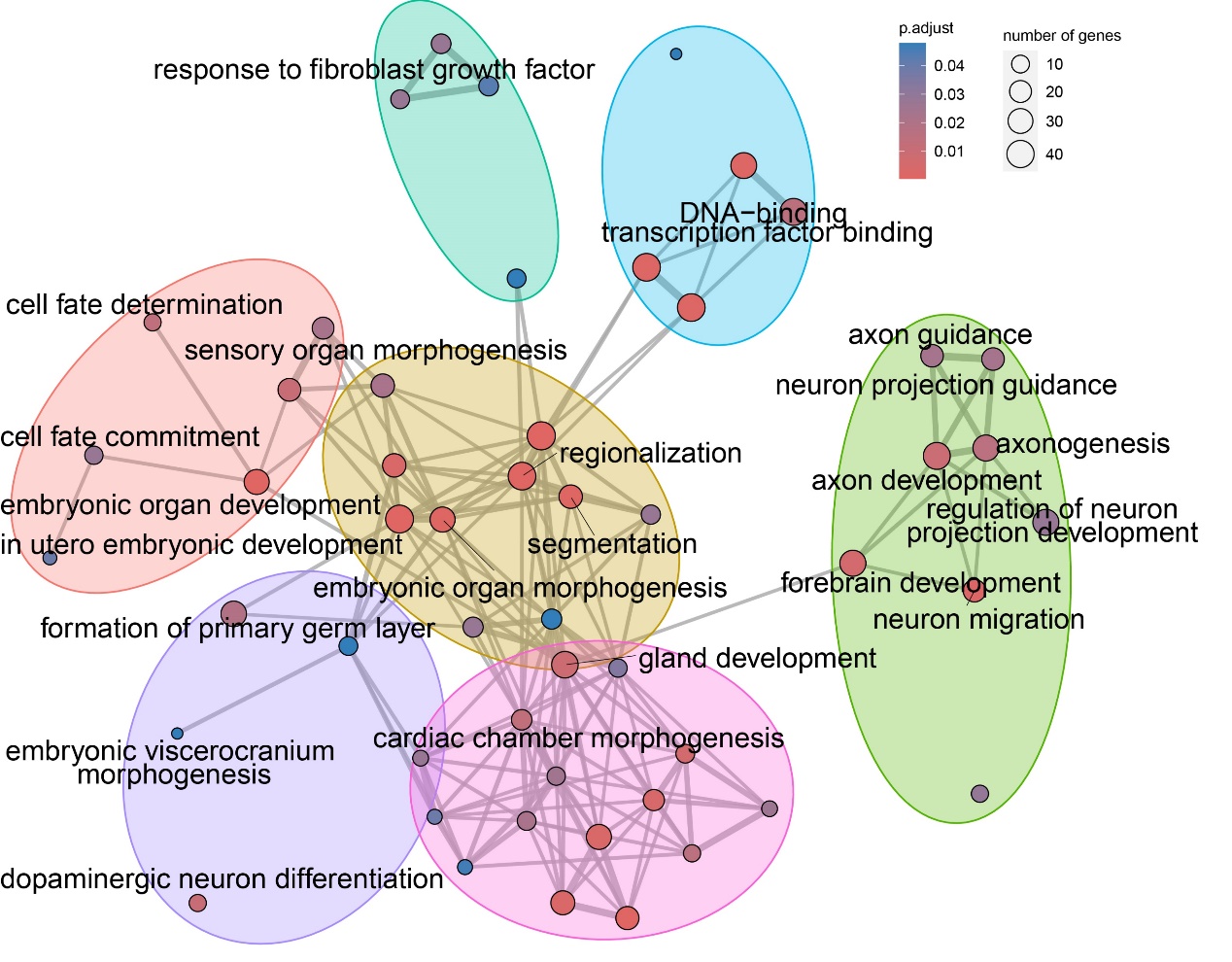

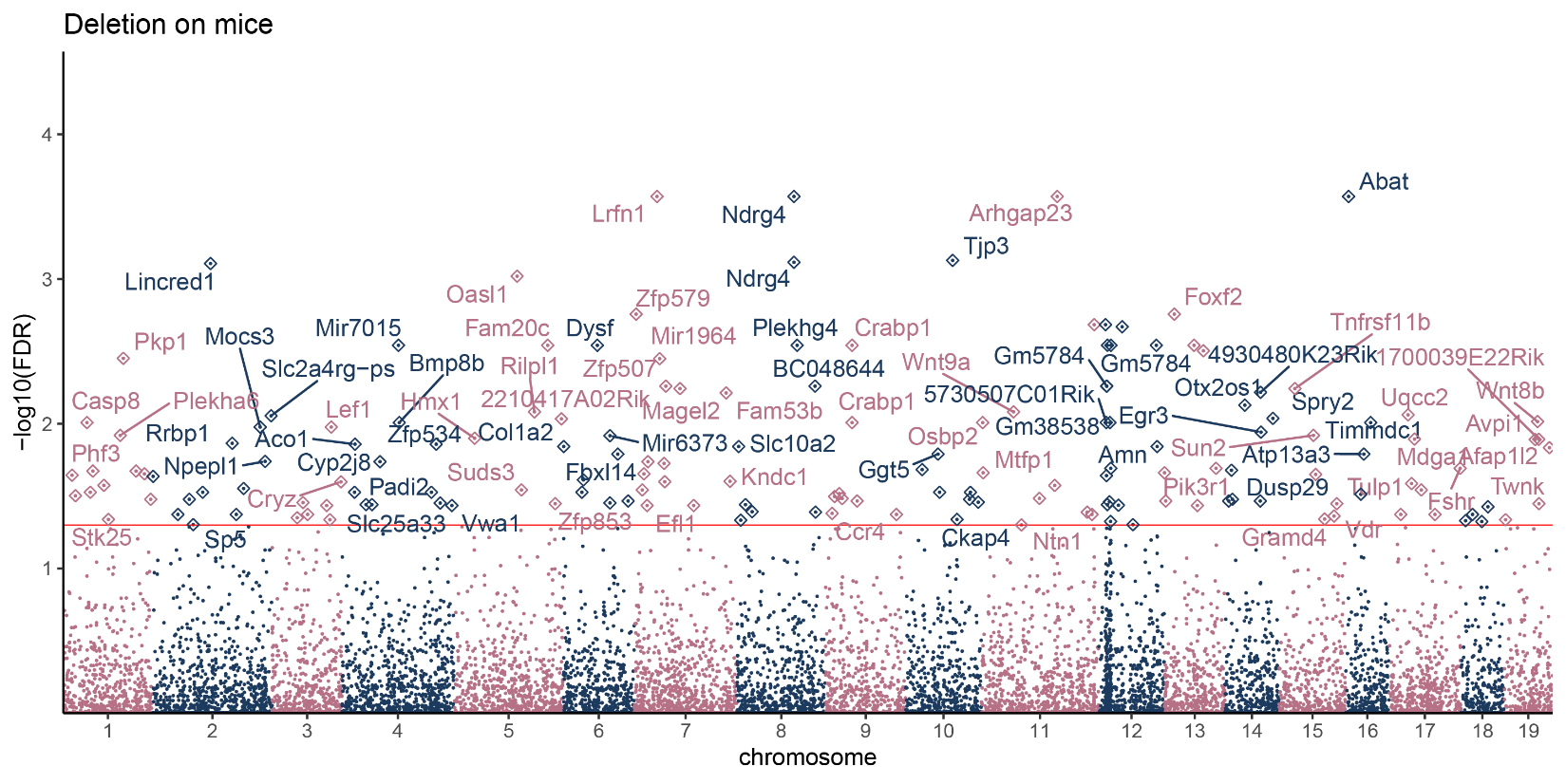
**Figure S3. Manhattan plot of deletion effects on mice.** Gene names were marked when they carried at least one DMP with FDR<0.05. The red line corresponds to the significant threshold.

**Figure S4 Gene Ontology enrichment based on DMPs between Tg and WT mice**

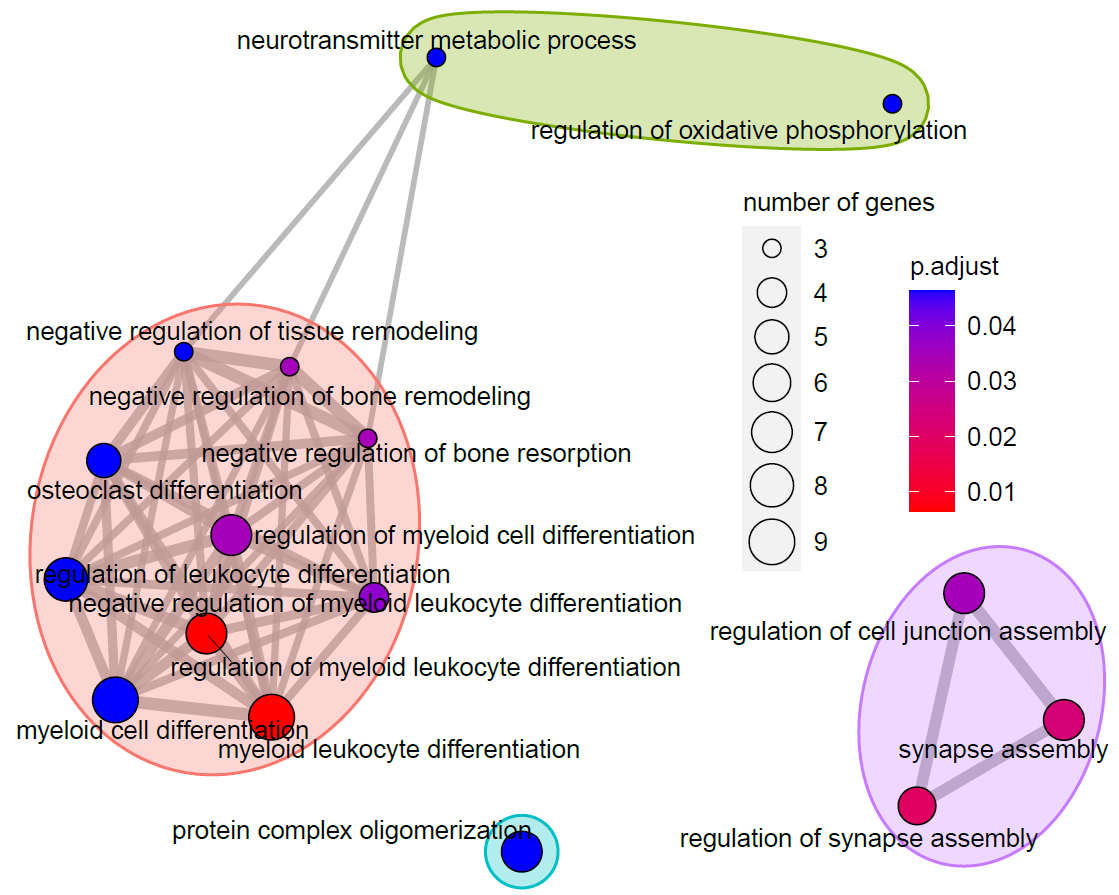

**Figure S5 miRNA expression comparison.** Volcano plot of miRNA expression differences identified in the comparison between individuals with 22q11.2DS and non-carriers (left) and in the comparison between individuals with 22q11.2DS and with/without SCZ (right).

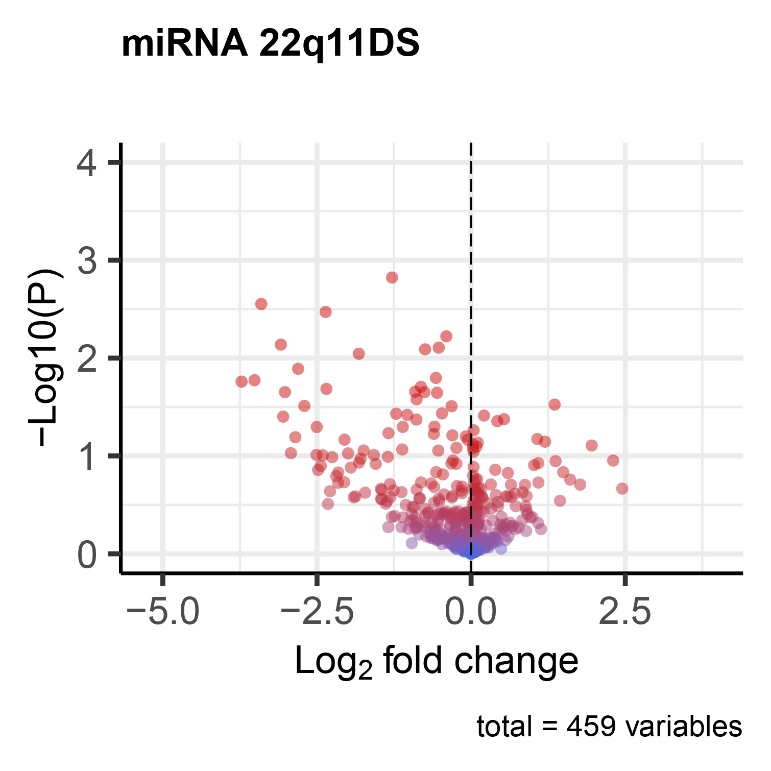

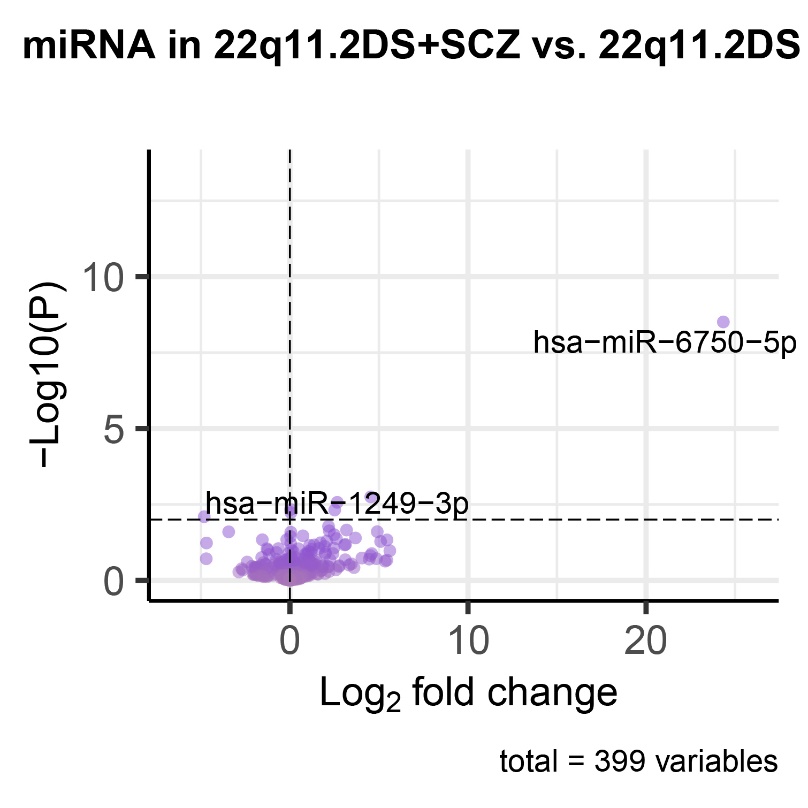

**Figure S6 : GO terms enrichment analysis from genes targeted by the miRNAs differentially expressed between individuals with 22q11.2DS and non-carriers**

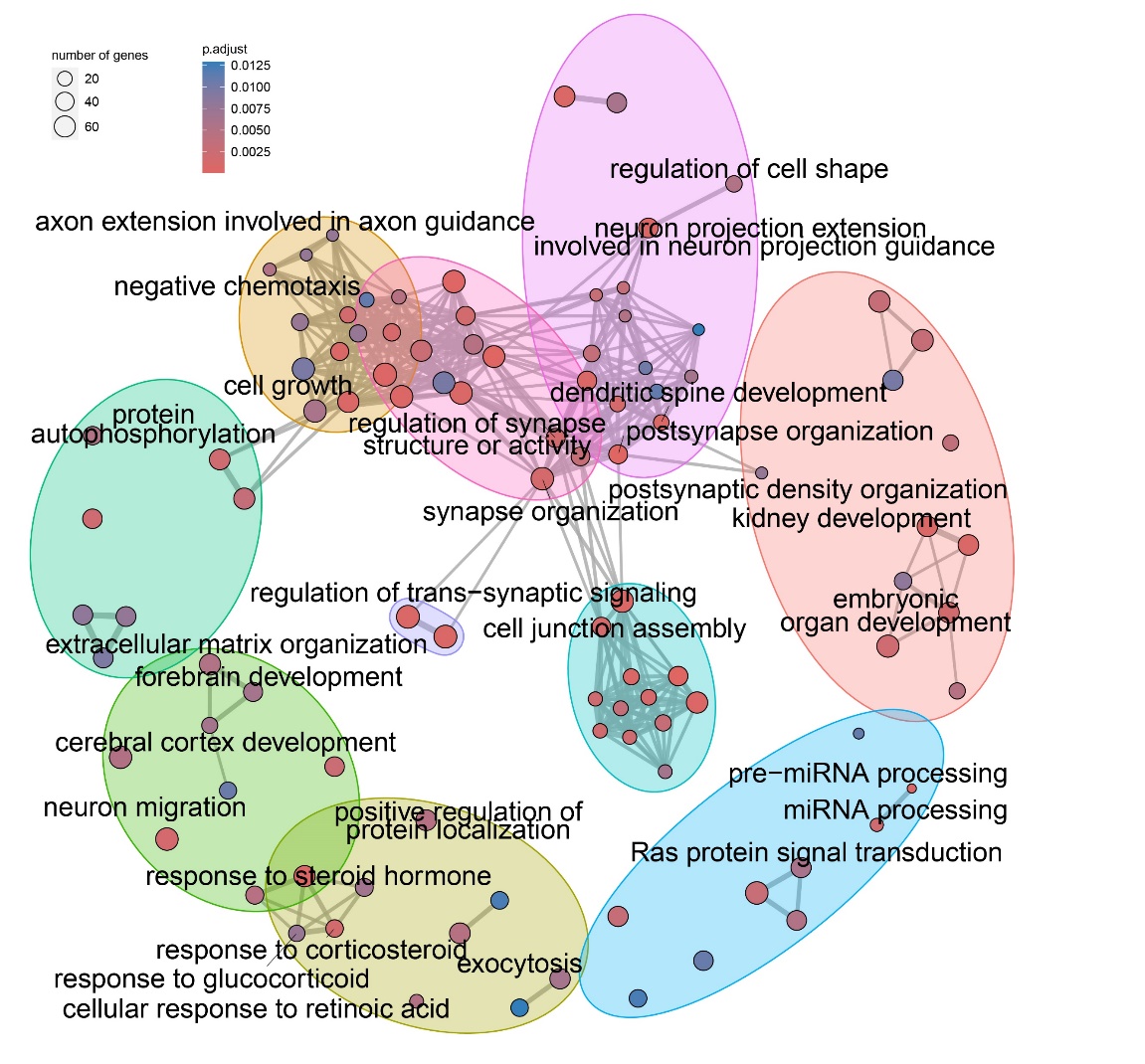

**Figure S7 : GO terms enrichment analysis from genes targeted by miRNAs differentially expressed between individuals with 22q11.2DS with and without SCZ**

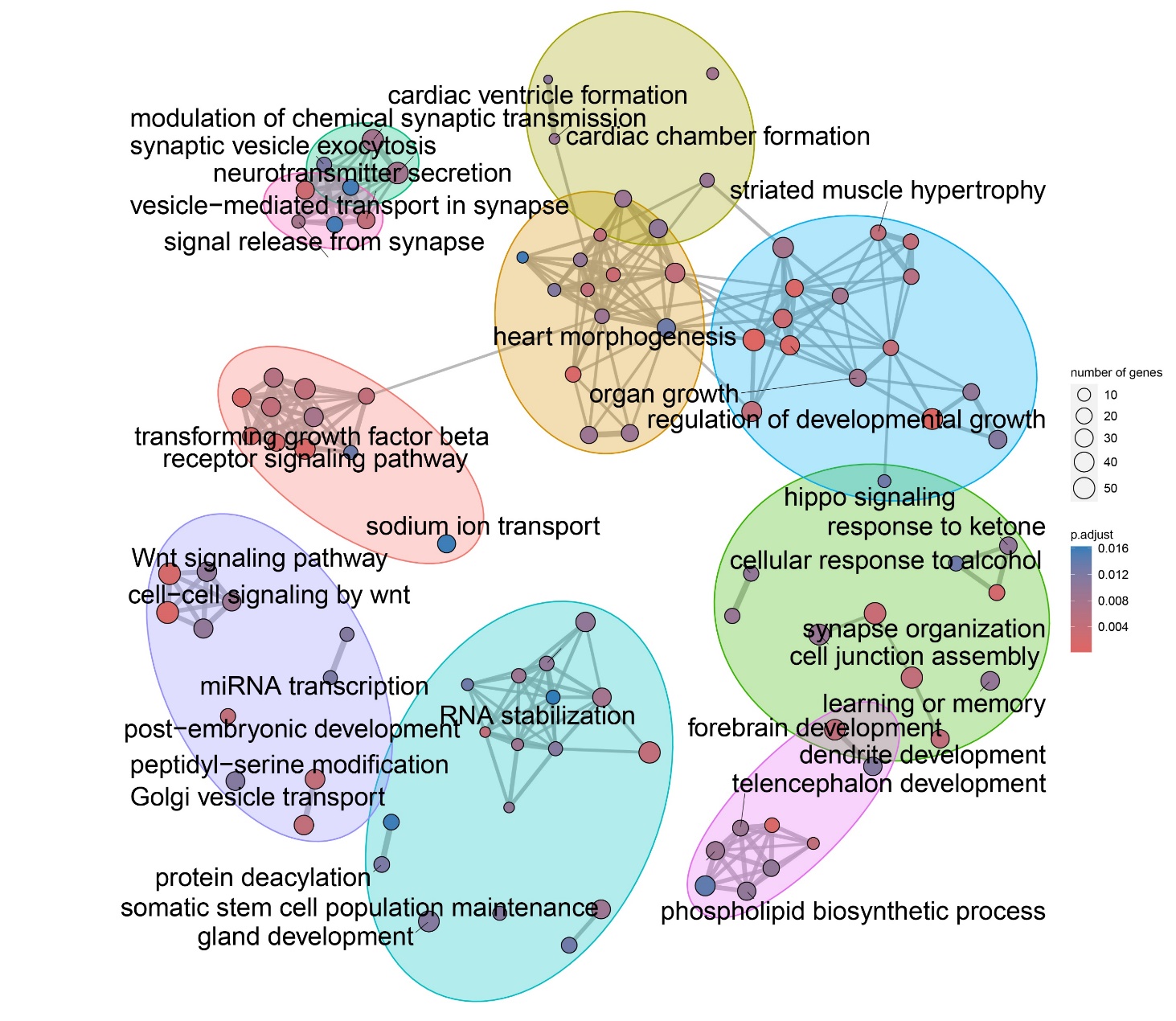

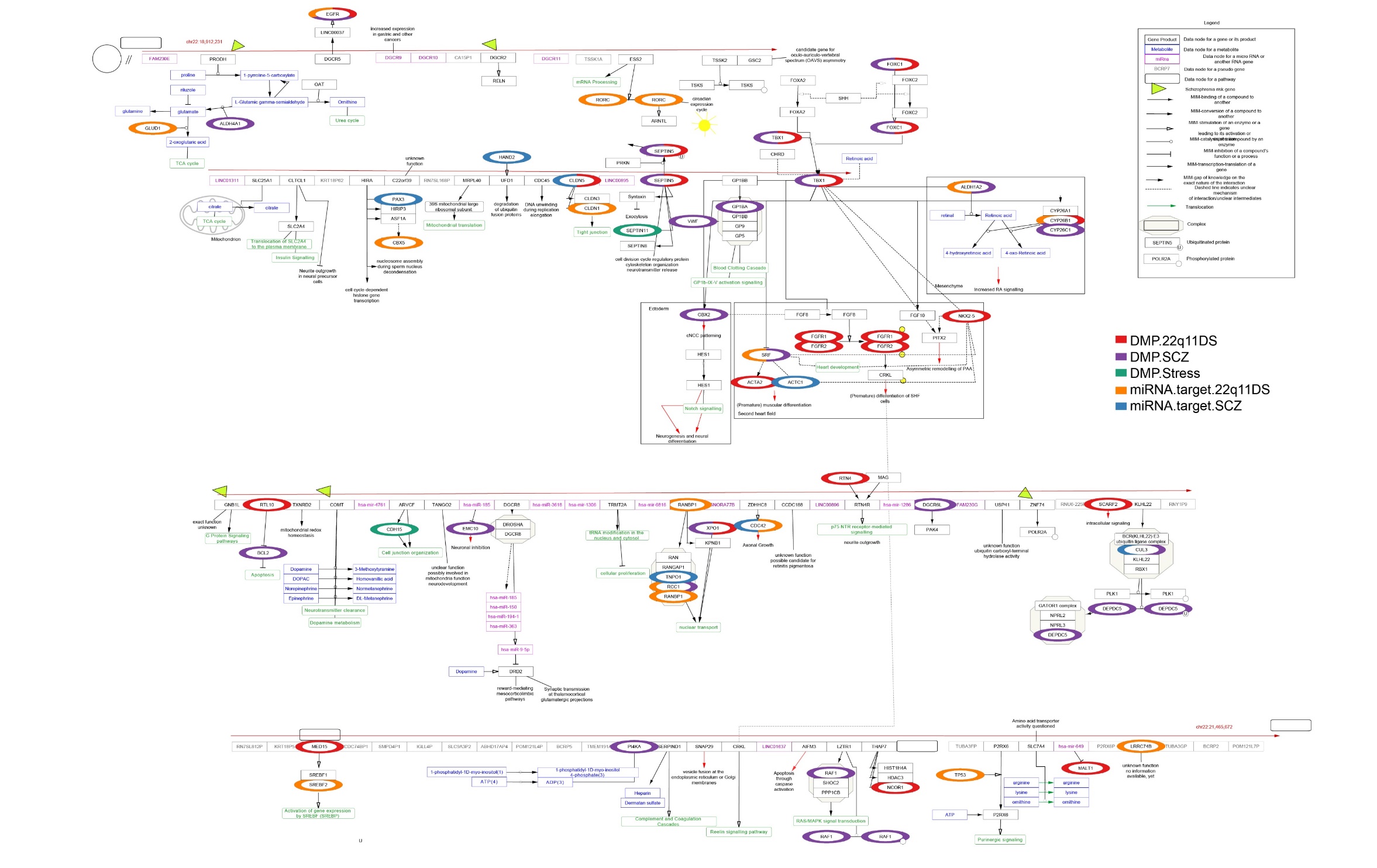
**Figure S8.** Summary of the findings in light of the 22q11 DS Wikipathway. Significant genes identified in each comparisons are highlighted in different colors (see legend)
